# Shedding light on the dynamic interplay of positive and negative symptoms of psychosis with Behavioral Tractography

**DOI:** 10.1101/2025.07.11.25331379

**Authors:** Andrea Imparato, Natacha Reich, Clémence Feller, Laura Ilen, Stephan Eliez, Christopher Graser, Maude Schneider, Corrado Sandini

**Author notes:** Corresponding author: Andrea Imparato, Developmental Imaging and Psychopathology Laboratory, Department of Psychiatry, University of Geneva School of Medicine, Campus Biotech, Chemin des Mines 9, 1202 Geneva, Switzerland. Co-Senior Authors (Maude Schneider, Corrado Sandini).

## Abstract

Our understanding of the interplay between positive and negative psychosis symptoms is constrained by reliance on retrospective assessments, which fail to capture dynamic, short-term symptom-context interactions. Ecological-Momentary-Assessment (EMA) offers real-time symptom tracking in naturalistic settings, but its validity and clinical utility remain uncertain. We address this using a recently developed Behavioral-Tractography approach in individuals with 22q11.2 Deletion Syndrome, a high-risk group for psychosis. Through Network-Dimensionality-Reduction, we found that dynamic EMA patterns closely mirror positive/negative symptom structure derived from gold-standard interviews, suggesting shared latent mechanisms across and within individuals and validating EMA’s assessment of dynamic psychosis symptom processes. Behavioral-Tractography provided informative dissection of symptom interactions—particularly between psychotic symptoms and motivational deficits—modulated by psychosis severity and distinguishing predominantly positive-vs-negative profiles. Vulnerability to psychological consequences varied independently of average symptom intensity, supporting the added value of EMA and Behavioral-Tractography for complementing clinical assessments and offering a novel lens on psychosis pathophysiology.

## Introduction

Psychosis is a complex psychiatric disorder characterized by multiple symptom dimensions, each with distinct phenomenology and treatment responses, reflecting different underlying pathophysiologies[1]. Still, these symptoms co-occur frequently enough to be considered manifestations of a single disorder[2]. Positive psychotic symptoms, such as delusions and hallucinations, are hallmarks of psychosis which respond effectively to antipsychotic medications targeting dysregulated dopaminergic activity in the mesolimbic pathway[3]. Negative symptoms, on the other hand, can be subdivided into two broad categories: (1) reduced motivation, including avolition and social withdrawal, and (2) diminished emotional and communicative expressiveness[4–6]. The pathophysiology of negative symptoms is less well understood and not entirely overlapping with that of positive symptoms[4, 7], as indicated by their less predictable and often limited response to antipsychotic treatment[8]. A distinction has been proposed between negative symptoms considered secondary consequence of primary positive symptoms (e.g., social isolation stemming from persecutory delusions) which improve with antipsychotic treatment, and primary negative symptoms, which are refractory to such treatments [9]. Such primary negative symptoms may stem from reduced dopaminergic transmission in the mesocortical pathway [10], potentially accounting for the paradoxical worsening of motivational deficits sometimes associated with high-dose antipsychotic medication[8, 9, 11]. Distinguishing primary from secondary negative symptoms however, currently depends largely on retrospectively observed treatment responses, highlighting a critical gap in our ability to tailor interventions effectively[5, 9, 12, 13].

This lack of knowledge may partially stem from the retrospective nature of psychiatric assessments, which has limited clinical descriptions of psychosis to a temporal resolution of weeks to months [13]. The dynamic relationship between short-term fluctuations in different clinical dimensions and environmental factors occurring over hours or days[14–16], could however be critical in mediating the pathophysiological interplay between positive and negative symptoms of psychosis [14, 17–24]. The recent advent of ambulatory assessment techniques, such as ecological momentary assessment (EMA), has enabled prospective and dynamical measurements of symptoms in the environmental context in which they occur, through questionnaires administered on smartphones multiple times a day, providing a unique window into such dynamic clinical phenomena[16, 25]. In this paper, we explore whether a recently developed Behavioral-Tractography approach could help exploit EMA’s full potential in advancing our understanding of the dynamic interplay between different psychosis symptom dimensions.

The first objective of the present study was to investigate whether EMA questionnaires can provide a sufficiently congruent representation of the complex clinical phenomena characterized with gold-standard psychosis assessments[14]. Similarity in psychometric instruments is typically established based on coherent low-dimensional structure, which is considered a reflection of their ability to capture similar underlying latent phenomena[26]. However, traditional dimensionality reduction techniques, which have been utilized to dissect the low-dimensional structure of positive versus negative symptoms [6], are not directly applicable to dynamically structured EMA data[25, 27, 28]. It thus remains fundamentally unclear whether similar phenomena are responsible for across-subject variation and within-subject dynamic fluctuations in positive and negative symptom intensity [27, 28]. We recently demonstrated that Network Dimensionality Reduction [29] can characterize the low-dimensional structure of dynamically fluctuating EMA behavioral data, with high consistency across independent samples of typically developing individuals, as well as in youth at high genetic vulnerability for psychosis due to 22q11.2 Deletion Syndrome [30–32]. Specifically, we identified a first rationality dimension that differentiated states of Affective Distress (e.g., *Anxiety, Sadness, Irritation*) from Psychotic-like Experiences (e.g., *Hallucinations, Feeling-Mistrustful, Confusing-Reality-with-Imagination*), which we defined as Cognitive-Distress[32]. A second valence dimension differentiated psychological distress from reversely coded wellbeing states, which were also segregated according to an affective to cognitive dimension, into Lacking-Affective-Wellbeing (e.g., *Lacking-Happiness, Lacking-Relaxation*) and Lacking-Cognitive-Wellbeing (*Lacking-Concentration, Lacking-Motivation, Lacking-Enjoyment*) [32]. Notably similar valence and rationality axes have been observed in studies of mental state attribution in others [33–37], suggesting that one’s understanding of others’ mental states may be informed by personal experience of psychological fluctuations [38]. Here we investigated whether the low-dimensional structure of dynamic EMA fluctuations would mirror variation in psychosis phenomenology measured with gold-standard clinical interviews across 22q11DS individuals. Such overlap would suggest similar latent pathophysiology underlying across-subject and within-subject variation in psychosis phenomenology [28] and represent an important step toward validating EMA measurement of dynamic psychosis symptom fluctuations [14].

The second objective of the present study was to shed light on dynamic behavioral pathways linking positive and negative symptoms at short time frames in everyday life. Indeed, beyond demonstrating their validity, the added value of EMA psychosis assessments also critically depends on deriving a sufficiently meaningful characterization of dynamic interactions between symptomatic dimensions and contextual factors, from highly complex time-dependent and multidimensional EMA data [39, 40]. In this regard, we recently proposed a novel Behavioral-Tractography analysis pipeline, providing a unique characterization of the broad architecture of behavioral dynamics, combined with a micro-scale characterization of the role of individual EMA behavioral variables [32]. This multi-scale view of behavioral dynamics proved clinically relevant as it differentiated individuals with 22q11DS from those with idiopathic autism spectrum disorder (ASD). In particular, 22q11DS individuals were vulnerable to the development of secondary motivational/cognitive wellbeing deficits, following the experience of social rejection associated with previous psychotic-like cognitive distress. Interestingly, this pathway mirrors the concept of secondary negative symptoms, but at a much faster temporal scale than previously anticipated[41]. Here we investigated whether Behavioral-Tractography could help dissect behavioral dynamics underlying variation in positive versus negative symptom dimensions, evaluated with gold-standard clinical instruments in individuals with 22q11DS. We hypothesized that Behavioral-Tractography would provide significant additional insights into the dynamic interplay between positive and negative symptoms of psychosis, unfolding in daily life. By providing a more intuitive characterization of behavioral dynamics, we propose that Behavioral-Tractography could assist in translating insights generated from EMA to improve psychosis clinical assessments.

## Results

### Clinical patterns detected by SIPS dimensionality reduction

Leveraging Principal Component Analysis (PCA) to address the inherent high dimensionality of SIPS data, we extracted two informative principal components (PCs) explaining a substantial portion of the variance (SIPS-Dimension-1: 42.9%, SIPS-Dimension-2: 10.3%) (Figure-1-Panel-1A).

**Figure 1:**
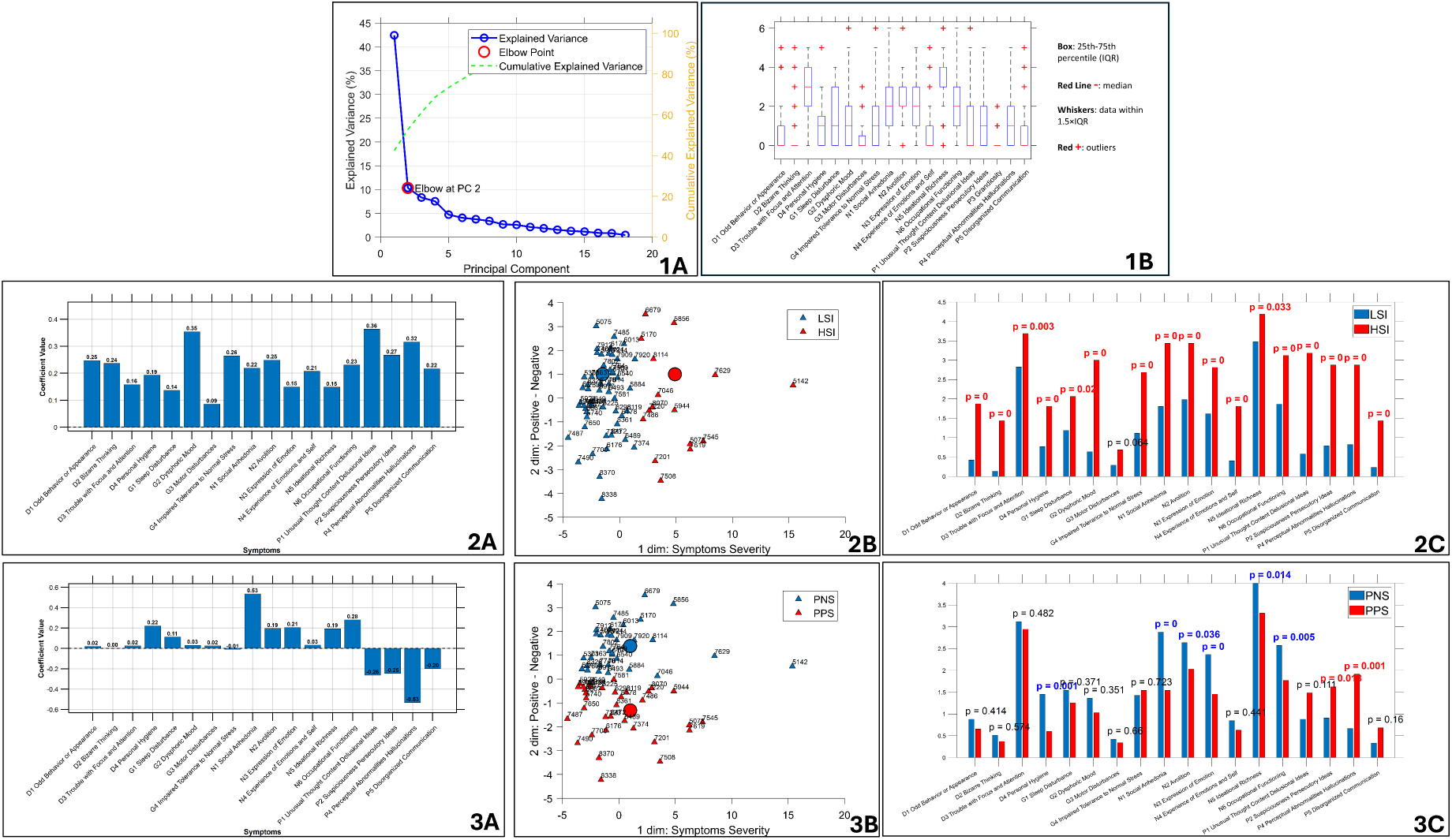
Results of PCA Dimensionality Reduction on SIPS Item scores characterizing main psychosis symptom dimensions. **Panel 1A:** Cumulative Explained Variance and Elbow Point in Principal Component Analysis The blue line represents the explained variance for each individual principal component (PC), while the green line shows the cumulative explained variance as PCs are successively added. The elbow point suggests an optimal number of PCs to retain. The x-axis denotes the number of PCs, and the y-axis represents the proportion of variance explained. **Panel 1B:** Distribution of SIPS Item Scores. Box plots illustrate the distribution of scores for each item on the Structured Interview for Prodromal Syndromes (SIPS). The x-axis represents individual SIPS items, while the y-axis shows the score range (0-6). **Panel 2A:** Bar plot showing the loadings of the first dimension resulting from Principal Component Analysis (PCA) performed on scores from the Structured Interview for Prodromal Syndromes (SIPS). The x-axis represents individual SIPS items, while the y-axis displays their corresponding loading values. SIPS-Dimension-1 captured overall symptom severity, as all SIPS items loaded positively along this dimension. **Panel 2B:** Scatter plot of subject loadings on the first two principal components derived from PCA performed on SIPS scores. The x-axis represents loadings on the first principal component (overall symptom severity), while the y-axis shows loadings on the second principal component (distinction between positive and negative symptoms). Each point represents an individual subject. Color coding reflects cluster membership determined by k-means clustering applied to the first principal component loadings. Blue points indicate subjects with lower overall symptom severity, while red points represent subjects with higher overall symptom severity. **Panel 2C:** Bar graph comparing average intensity of SIPS items between High-Symptom-Intensity subjects (red) and Low-Symptom-Intensity subjects (blue). The x-axis displays individual SIPS items, while the y-axis shows the mean score for each item. **Panel 3a:** Bar plot showing the loadings of the second dimension resulting from Principal Component Analysis (PCA) performed on scores from the Structured Interview for Prodromal Syndromes (SIPS). The x-axis represents individual SIPS items, while the y-axis displays their corresponding loading values. SIPS-Dimension-2 captures the distinction between positive and negative symptoms, with positive symptoms loading negatively and negative symptoms loading positively **Panel 3B:** Scatter plot of subject loadings on the first two principal components derived from PCA performed on SIPS scores. The x-axis represents loadings on SIPS-Dimension-1 (overall symptom severity), while the y-axis shows loadings on SIPS-Dimension-2 (distinction between positive and negative symptoms). Each point represents an individual subject. Color coding reflects cluster membership determined by k-means clustering applied solely to the loadings on SIPS-Dimension-2. Blue points indicate subjects with a predominantly negative symptoms (PNS), while red points represent subjects with a predominantly positive symptoms (PPS). **Panel 3C:** Bar graph comparing average intensity of SIPS items between predominant-negative and predominant-positive subjects. The x-axis displays individual SIPS items, while the y-axis shows the mean score for each item.

SIPS-Dimension-1 captured overall psychotic symptom severity as every positive, negative, disorganization and general SIPS item loaded positively along this dimension, with strongest contributions for positive symptoms such as *Unusual-Thought-Content*, *Perceptual-Abnormalities, Persecutory-Ideas* and for the general *Dysphoric-Mood* item (Figure-1-Panel-2A). K-means clustering on SIPS-Dimension-1 scores separated a Low Symptom Intensity subgroup (LSI; n=53), with lower SIPS-Dimension-1 scores, from a High Symptom Intensity subgroup (HSI; n=15) ( Figure-1-Panels-2B). Compared to LSI the HSI presented higher intensity of all SIPS items except for *Motor-Disturbances* (Figure-1-Panels-2C).

SIPS-Dimension-2 captured the distinction between positive symptoms, which had positive loading, and negative symptoms, which loaded negatively (Figure-1-Panels-3A). K-means clustering on PC2 scores separated a first Predominantly Positive Symptoms subgroup (PPS; n=31) with higher SIPS-Dimension-2 scores, displaying significantly higher intensity for some positive symptoms (*Hallucinations* and *Persecutory-Ideas*), from a second Predominantly Negative Symptoms subgroup (PNS; n=37), with lower SIPS-Dimension-2 scores, characterized by higher intensity in all negative symptoms except for *Experience-of-Emotions* as well as higher *Impaired-Personal-Hygiene* disorganization item (Figure-1-Panels-3B-C).

There were no significant differences in demographic information (age and gender) between subjects within either cluster pair (HSI vs. LSI and PPS vs. PNS; See Table 1 for details). Full-Scale IQ was higher in LSI compared to HSI (LSI=75,65+/-13,07, HSI=65,40+/-13,45, p=0.01) while it did not differ across PPS and PNS subgroups (PPS=73,22+/-12,17, PNS= 73,19+/-15,46, p=0.99). This two-dimensional framework based on PCA-derived SIPS dimensions, which were largely consistent with previous reports [42, 43], provided a basis to investigate the relationships between SIPS symptom patterns and EMA variables.

**Table 1:** Demographic information for the 4 populations. Gender differences were tested with Chi-Square test. Continuous variables were tested with Two-Sample T-Tests. Full Scale IQ differed between LSI and HSI subgroups.

| **Demographic Table** | **LSI**<br>**(N=52)** | **HSI**<br>**(N=16)** | **P-value HSI-LSI** | **PNS**<br>**(N=33)** | **PPS**<br>**(N=35)** | **P-value PPS-PNS** | **Full Sample**<br>**(N=68)** |
| --- | --- | --- | --- | --- | --- | --- | --- |
| **Age:** mean (sd) | 19.48 (5.17) | 18.88 (5.36) | 0.69 | 20.09 (5.37) | 18.62 (4.97) | 0.24 | 19.34 (5.18) |
| **Gender:** M/F | 27/25 | 10/6 | 0.47 | 21/12 | 16/19 | 0.14 | 37/31 |
| **Assessments:**<br>mean (sd) | 25.62 (11.23) | 25.06 (9.92) | 0.86 | 26.33 (11.53) | 24.69 (10.3) | 0.54 | 25.49(10.86) |
| **Full Scale IQ:**<br>mean (sd) | 75.65 (13.07) | 65.40 (13.45) | **0.01** | 73,19 (15.46) | 73.22 (12.17) | 0.99 | 73.20(13.77) |

### Correlation between SIPS clinical pattern and EMA symptom intensity

SIPS-Dimension-1, reflecting overall psychotic symptom severity, demonstrated strong positive correlations with the average intensity of all EMA items related to positive psychotic symptoms, such as *Hallucinations* (R=-0.49, p<0.001), *Feeling-Unsafe* (R=-0.57, p<0.001), and *Confusing-Reality-with-Imagination* (R=-0.42, p<0.001) and a significant while weaker associations with all affective distress variables such as *Sadness* (R=-0.35, p<0.001)*, Irritation* (R=-0.33, p<0.01)*, Anxiety* (R=-0.26, p<0.03)*, Feeling-Rejected* (R=-0.41, p<0.001) (Table 2). Indeed, the average intensity of all such affective and cognitive distress variables, as well as *Fatigue, Lacking-Physical-Activity and Lacking-Confidence* (all p<0.001) were significantly increased in the HSI compared to LSI, who instead reported higher *Lacking-Concentration* (p<0.001)*, Lacking-Motivation*(p<0.001)*, Lacking-Enjoyment*(p<0.001) and *Finding-Activity-Difficult* (p<0.02) (Table 2).

**Table 2.** Symptoms intensities measured via Ecological Momentary Assessment (EMA) across populations and their correlations with psychopathology dimensions derived from SIPS-PCA. Mean Intensity (LSI, HSI, PNS, PPS) and Std Intensity (LSI, HSI, PNS, PPS) report the average and standard deviation of EMA symptom intensity scores for Low Symptom Intensity (LSI), High Symptom Intensity (HSI), Predominant Negative Symptoms (PNS), and Predominant Positive Symptoms (PPS) populations. p-value (HSI-LSI, PPS-PNS) displays the results of independent two-sample t-tests comparing mean symptom intensities between LSI and HSI populations, as well as between PPS and PNS subgroups. Columns R and p-value represent the Pearson correlation coefficients and significance levels for the relationships with SIPS-PCA Dimension 1 (High – Low intensity) and Dimension 2 (Positive - Negative symptoms).

| EMA symptoms | High-/ Low-Symptom-Intensity Dimension |  |  |  |  |  |  | Predominant Positive- negative Symptom Dimension |  |  |  |  |  |  |
| --- | --- | --- | --- | --- | --- | --- | --- | --- | --- | --- | --- | --- | --- | --- |
|  | Mean Intensity LSI | Std Intensity LSI | Mean Intensity HSI | Std Intensity HSI | p-value HSI-LSI | R | p-value | Mean Intensity PNS | Std Intensity PNS | Mean Intensity PPS | Std Intensity PPS | p-value PPS-PNS | R | p-value |
| Lack Relaxation | 3.26 | 1.55 | 3.22 | 1.72 | 0.71 | 0.00 | 0.98 | 3.40 | 1.57 | 3.10 | 1.60 | **0.00** | 0.12 | 0.3 |
| Loneliness | 1.62 | 1.30 | 1.84 | 1.39 | **0.00** | 0.28 | **0.02** | 1.69 | 1.34 | 1.64 | 1.31 | 0.44 | -0.1 | 0.3 |
| Anxiety | 1.81 | 1.41 | 2.09 | 1.49 | **0.00** | 0.26 | **0.03** | 1.90 | 1.45 | 1.85 | 1.42 | 0.53 | -0.0 | 0.5 |
| Lack Happiness | 3.38 | 1.63 | 3.47 | 1.82 | 0.36 | 0.08 | 0.51 | 3.57 | 1.66 | 3.23 | 1.68 | **0.00** | 0.11 | 0.3 |
| Irritation | 1.45 | 1.12 | 1.84 | 1.48 | **0.00** | 0.33 | **0.01** | 1.51 | 1.21 | 1.57 | 1.24 | 0.27 | -0.1 | 0.1 |
| Sensory Issue | 1.23 | 0.78 | 1.80 | 1.25 | **0.00** | 0.53 | **0.00** | 1.39 | 1.02 | 1.34 | 0.86 | 0.30 | -0.0 | 0.8 |
| Lack Excitement | 5.28 | 1.94 | 5.30 | 2.09 | 0.89 | -0.16 | 0.19 | 5.29 | 1.97 | 5.28 | 1.99 | 0.90 | 0.10 | 0.4 |
| Sadness | 1.51 | 1.22 | 2.06 | 1.62 | **0.00** | 0.35 | **0.00** | 1.49 | 1.18 | 1.78 | 1.47 | **0.00** | -0.2 | **0.0** |
| Lack Confidence | 3.56 | 1.73 | 4.01 | 1.62 | **0.00** | 0.07 | 0.59 | 3.79 | 1.76 | 3.53 | 1.66 | **0.00** | 0.08 | 0.5 |
| Feeling Rejected | 1.32 | 0.92 | 1.99 | 1.56 | **0.00** | 0.41 | **0.00** | 1.47 | 1.12 | 1.47 | 1.14 | 0.99 | -0.1 | 0.2 |
| Feeling Unsafe | 1.29 | 0.94 | 2.40 | 1.91 | **0.00** | 0.57 | **0.00** | 1.45 | 1.16 | 1.65 | 1.46 | **0.00** | -0.1 | 0.3 |
| Confusion | 1.18 | 0.75 | 1.52 | 1.17 | **0.00** | 0.42 | **0.00** | 1.33 | 1.05 | 1.19 | 0.65 | **0.00** | 0.00 | 0.9 |
| Hallucinations | 1.21 | 0.81 | 2.18 | 1.75 | **0.00** | 0.49 | **0.00** | 1.33 | 1.05 | 1.53 | 1.28 | **0.00** | -0.1 | 0.3 |
| Feeling Tired | 3.09 | 1.85 | 3.63 | 1.56 | **0.00** | 0.23 | 0.06 | 3.06 | 1.83 | 3.37 | 1.77 | **0.00** | 0.00 | 1.0 |
| Lack Motivation | 4.41 | 1.76 | 4.03 | 2.07 | **0.00** | -0.15 | 0.23 | 4.45 | 1.74 | 4.20 | 1.93 | **0.00** | 0.10 | 0.4 |
| Lack Physical Activity | 4.77 | 1.95 | 5.16 | 1.75 | **0.00** | -0.06 | 0.60 | 4.94 | 1.88 | 4.78 | 1.94 | 0.08 | 0.04 | 0.7 |
| Finding Activity Difficult | 1.72 | 1.37 | 1.55 | 1.12 | **0.02** | 0.18 | 0.14 | 1.75 | 1.37 | 1.61 | 1.25 | **0.04** | 0.18 | 0.1 |
| Lack Enjoing Activity | 3.23 | 1.62 | 2.77 | 1.75 | **0.00** | -0.21 | 0.09 | 3.14 | 1.53 | 3.11 | 1.78 | 0.70 | -0.0 | 0.9 |
| Lack Concentration | 4.10 | 1.94 | 3.69 | 2.07 | **0.00** | -0.14 | 0.25 | 4.07 | 2.01 | 3.94 | 1.94 | 0.17 | 0.07 | 0.5 |
| Being Alone | 0.36 | 0.48 | 0.35 | 0.48 | 0.54 | 0.03 | 0.84 | 0.38 | 0.49 | 0.33 | 0.47 | **0.03** | -0.0 | 0.8 |

SIPS-Dimension-2, distinguishing negative and positive symptom dimensions, was negatively correlated with EMA *Sadness* (R=-0.23, p<0.001). *Sadness* was indeed significantly increased in the PPS subgroup, who also reported higher *Hallucinations, Feeling-Unsafe* and *Fatigue* (all p<0.001) compared to the PNS subgroup (Table 2). The PNS subgroup instead reported significantly higher intensity for most EMA items reflecting lack of well-being including *Lacking-Relaxation, Lacking-Happiness, Lacking-Confidence, Lacking-Motivation* (all p<0.001) as well as higher *Confusing-Reality-with-Imagination* (p<0.001) and higher social isolation (*Being-Alone, p=0.03)* compared to the PPS group (Table 2 and Figure-2-Panel-3A). Overall, this suggests that SIPS-Dimesion-2 differences of negative vs positive symptom dimensions across PNS and PPS subgroups, were mirrored by similar differences in average intensity of EMA variables measuring psychological distress vs lacking psychological wellbeing.

**Figure 2:**
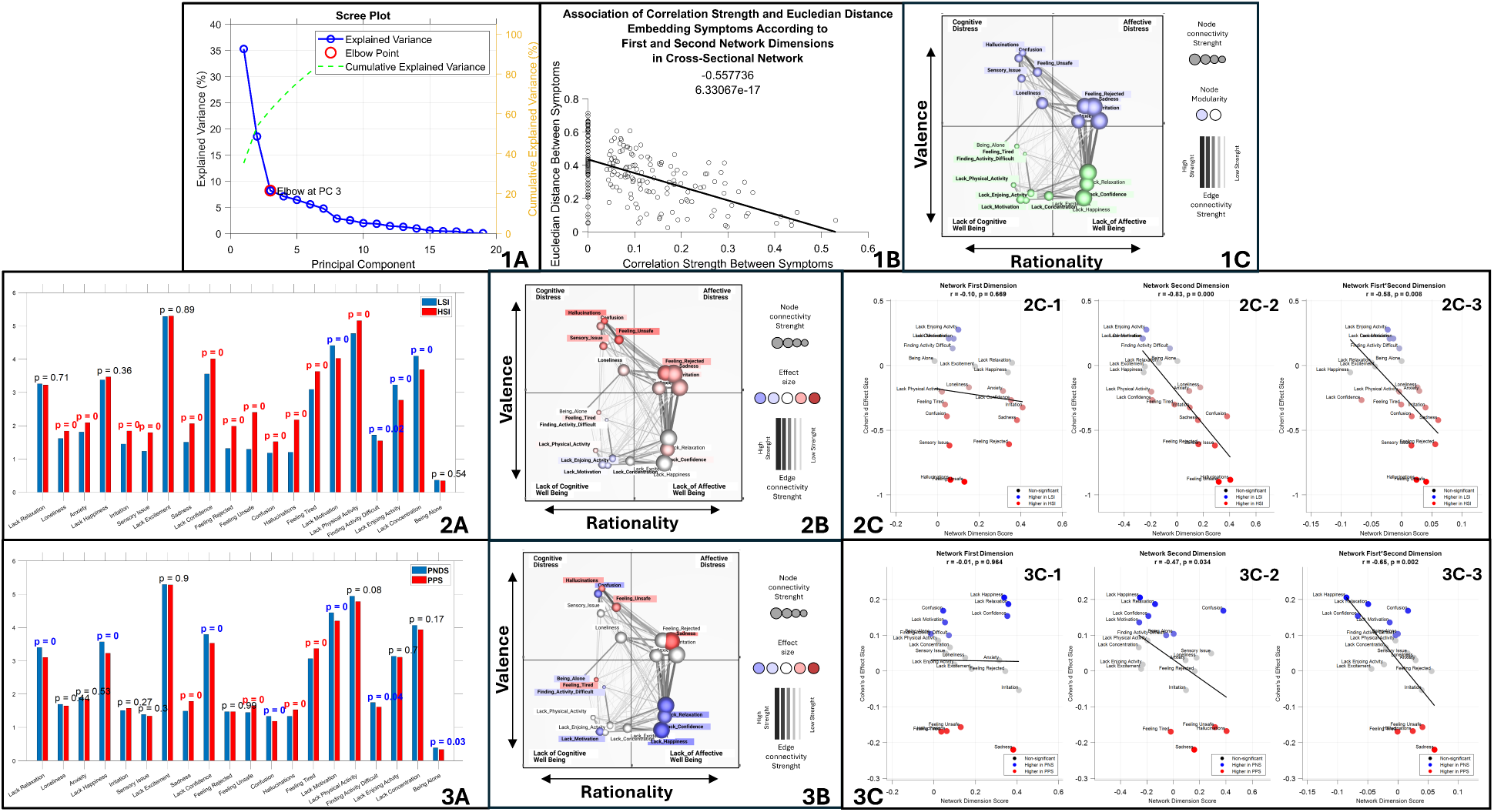
Low-Dimensional Structure of dynamic EMA fluctuations revealed by Network Dimensionality Reduction and association with across-subject variation in SIPS Psychosis symptom dimensions. **Panel 1A:** Principal Component Analysis (PCA) performed on the 2D-Network Adjacency Matrix capturing the association strength between dynamic fluctuations of EMA variables, after accounting for across-subject variation in average intensity with Subject-specific Random-Intercept Mixed-Models-Linear-Regression. The blue line represents the explained variance for each individual principal component (PC), while the green line shows the cumulative explained variance as PCs are successively added. The elbow point suggests an optimal number of PCs to retain. The x-axis denotes the number of PCs, and the y-axis represents the proportion of variance explained (0-1). **Panel 1B:** Association of Euclidian Distance between Variables derived from NDR and empirically measured correlation strength between variables, estimating overall accuracy of NDR representation. **Panel 1C:** Single-layer networks representing cross-sectional patterns of co-occurrence between psychological and contextual variables at specific moments in time in the entire 22q11DS sample. Each psychological and contextual variable is represented as an individual network node. Thickness and transparency of network edges represent the strength of cross-sectional statistical associations between variables. Node size represents the summed connectivity strength of the corresponding psychological or contextual variable. Node positions is determined according to two main dimensions derived from network dimensionality reduction (NDR) and capturing the differential propensity for variables to manifest together. The first NDR Dimension is represented along the horizontal dimension and differentiates variables according to *rationality:* Affective states cluster on the right while Cognitive states are clustered on the left. The second NDR Dimension is represented along the vertical dimension and differentiates variables according to *valence:* Psychological Distress states cluster in the upper part of the graph, while Lack of Psychological Wellbeing (reversely coded Wellbeing variables) cluster in the bottom part of the graph. **Panel 2A:** Bar graph comparing average intensity of EMA items between High-Symptom-Intensity subjects (red) and Low-Symptom-Intensity subjects(blue) defined according to SIPS-Dimension-1. The x-axis displays individual EMA items, while the y-axis shows the mean score for each item. Panel **2B:** Single-layer networks representing cross-sectional patterns of co-occurrence between psychological and contextual variables at specific moments in time. Each psychological and contextual variable is represented as an individual network node. Thickness and transparency of network edges represent the strength of cross-sectional statistical associations between variables. Node size represents the summed connectivity strength of the corresponding psychological or contextual variable. Node colors are scaled according to the effect size of the difference in intensity of the corresponding psychological or contextual variable between HSI and LSI. **Panel 2C:** Correlation EMA Intensity Differences Effect Sizes (Cohen’s d) in High-Symptom-Intensity vs Low-Symptom-Intensity subjects, capturing their association with SIPS-Dimension-1 and loading of such EMA items according to NDR dimensions, capturing the pattern with which they fluctuated dynamically with other EMA variables. The y-axis displays Cohen’s *d* effect sizes comparing EMA item intensity High-Symptom-Intensity vs Low-Symptom-Intensity subjects. Each point corresponds to an EMA item, colored by effect direction (red: higher intensity in HSI; blue: higher intensity in LSI; black: non-significant differences). A regression line illustrates the correlation between MLN coordinates and effect sizes. **Panel 2C1:** The x-axis represents coordinates from the first rationality NDR dimension (cognitive to affective). **Panel 2C-2:** The x-axis represents coordinates from the second valence NDR Dimensions (Lack of Well being to Distress). **Panel 2C-3:** The x-axis represents product of the first two coordinates of MLN. **Panel 3A:** Bar graph comparing average intensity of EMA items between Predominantly-Positive-Symptoms subjects (red) and Predominantly-Negative-Symptoms(blue) defined according to SIPS-Dimension-2. The x-axis displays individual EMA items, while the y-axis shows the mean score for each item. **Panel 3B:** Single-layer networks representing cross-sectional patterns of co-occurrence between psychological and contextual variables at specific moments in time. Each psychological and contextual variable is represented as an individual network node. Thickness and transparency of network edges represent the strength of cross-sectional statistical associations between variables. Node size represents the summed connectivity strength of the corresponding psychological or contextual variable. Node colors are scaled according to the effect size of the difference in intensity of the corresponding psychological or contextual variable between PPS and PNS. **Panel 3C:** Correlation EMA Intensity Differences Effect Sizes (Cohen’s d) in Predominantly-Positive-Symptoms vs Predominantly-Negative-Symptoms, capturing their association with SIPS-Dimension-2 and loading of such EMA items according to NDR dimensions, capturing the pattern with which they fluctuated dynamically with other EMA variables. The y-axis displays Cohen’s *d* effect sizes comparing EMA item intensity Predominantly-Positive-Symptoms vs Predominantly-Negative-Symptoms subjects. Each point corresponds to an EMA item, colored by effect direction (red: higher intensity in PPS; blue: higher intensity in PNS; black: non-significant differences). A regression line illustrates the correlation between MLN coordinates and effect sizes. **Panel 3C1:** The x-axis represents coordinates from the first rationality NDR dimensions (cognitive to affective). **Panel 3C-2:** The x-axis represents coordinates from the second dimension valence NDR Dimensions (Lack of Well being to Distress). **Panel 3C-3:** The x-axis represents product of the first two coordinates of MLN.

### Low-Dimensional structure of dynamic EMA fluctuations and correspondence with SIPS Dimension

In accordance with previous findings [32], only the first 2 dimensions derived from Network Dimensionality Reduction (NDR), accounting for 35% and 18% of variance in the network structure, meaningfully characterized the propensity for EMA variables to co-occur, as estimated by a significant negative correlation between Euclidian distance separating variables, and empirically observed association strength (R=-0.55, p<0.001) (Figure-2-Panel-1B). Consistently with previous literature [32], the first NDR Dimension differentiated affective from cognitive variables along a right-to-left horizontal *Rationality* axis, while the second dimension differentiated psychological distress from lacking psychological wellbeing along a upper-to-lower vertical valence axis. Embedding EMA variables according to such dimensions hence resulted in four network quadrants (Figure-2-Panel-1C):

- Upper-Right: Affective-Distress (*Sadness, Irritation, Anxiety, Feeling-Rejected*).
- Lower-Right: Lacking-Affective-Well-Being (*Lacking-Happiness, Lacking-Relaxation, Lacking-Confidence*).
-Lower-Left: Lacking-Cognitive-Wellbeing (*Lacking-Motivation, Lacking-Concentration, Lacking-Enjoyment*).
-Upper-Left: Cognitive/Psychotic-Distress (*Hallucinations*, *Feeling-Unsafe, Confusing-Reality-with-Imagination)*.

*Loneliness* had an intermediate position in the upper portion of the network, linking social isolation (*Being-Alone*) in the lower-left Lacking-Cognitive-Wellbeing quadrant to upper-left Cognitive-Distress and upper-right Affective-Distress variables.

Interestingly, NDR Dimensions capturing the propensity for EMA variables to fluctuate together also tightly recapitulated the pattern of correlation between the average intensity of EMA variables and SIPS dimensions. Specifically, the strength of SIPS-Dimensions-1-EMA association was significantly anti-correlated with loading of EMA-items along the NDR valence dimension (R=-0.83, p<0.001) (Figure-2-Panel-2C-2). As such EMA items reported with higher intensity in the HSI compared to the LSI subgroup, preferentially clustered in the upper psychological distress portion of the EMA-Network, with particularly strong differences for Psychotic/Cognitive-Distress items clustering in the upper-left network quadrant (Figure-2-Panel-2B).

SIPS-Dimension-2 capturing the differential Negative-vs-Positive symptom severity also showed associations with EMA-NDR dimensions, which were particularly strong for the product of Valence and Rationality dimensions (R=0.65, p<0.001) (Figure-2-Panel-3C-3). As such, EMA items that were increased in the PPS subgroup, clustered preferentially in the upper-right Affective-Distress EMA quadrant, including in particular *Sadness,* as well as *Hallucinations, Feeling-Unsafe and Feeling-Tired,* clustering in the upper-right portions of their respective Cognitive-Distress and Lacking-Cognitive-Wellbeing quadrants. EMA items that were increased in PNS individuals instead clustered preferentially in the lower-left Lacking-Cognitive-Wellbeing variables (*Lacking-Motivation, Being-Alone, Finding-Activity-Difficult*) and lower-right Lacking-Affective-Wellbeing variables (*Lacking-Happiness, Lacking-Relaxation, Lacking-Confidence*). The only psychological distress variable to be increased in the PNS subgroup was *Confusing-Reality-with-Imagination,* located in the leftmost portion of the Cognitive-Distress quadrant (Figure-2-Panel-3B).

Such correspondence in low-dimensional structure would suggest that latent phenomena underpinning variation in SIPS positive vs negative symptom intensity across subjects may be linked to mechanisms underpinning short term dynamic fluctuation of EMA symptom intensity, reported by individual participants, from one assessment to the next. This would support the validity of EMA for measuring fluctuations in the same positive and negative symptom dimensions that are measured retrospectively with current gold-standard clinical instruments. We, therefore, next explored whether EMA could provide additional insights on dynamic interplay between symptom dimensions in the flow of daily life.

### Association between SIPS-PCA Dimension and EMA Dynamic Network Structure

We firstly investigated how SIPS-PCA dimensions modulated the strength of individual associations between pairs of EMA variables. In the 3D-MLN view, cross-sectional connections that were significantly modulated by SIPS-PCA dimensions were represented in cross-sectional temporal layers, while time-lagged connections connected temporal layers along the left to right temporal axis (Figure-3).

**Figure 3.**
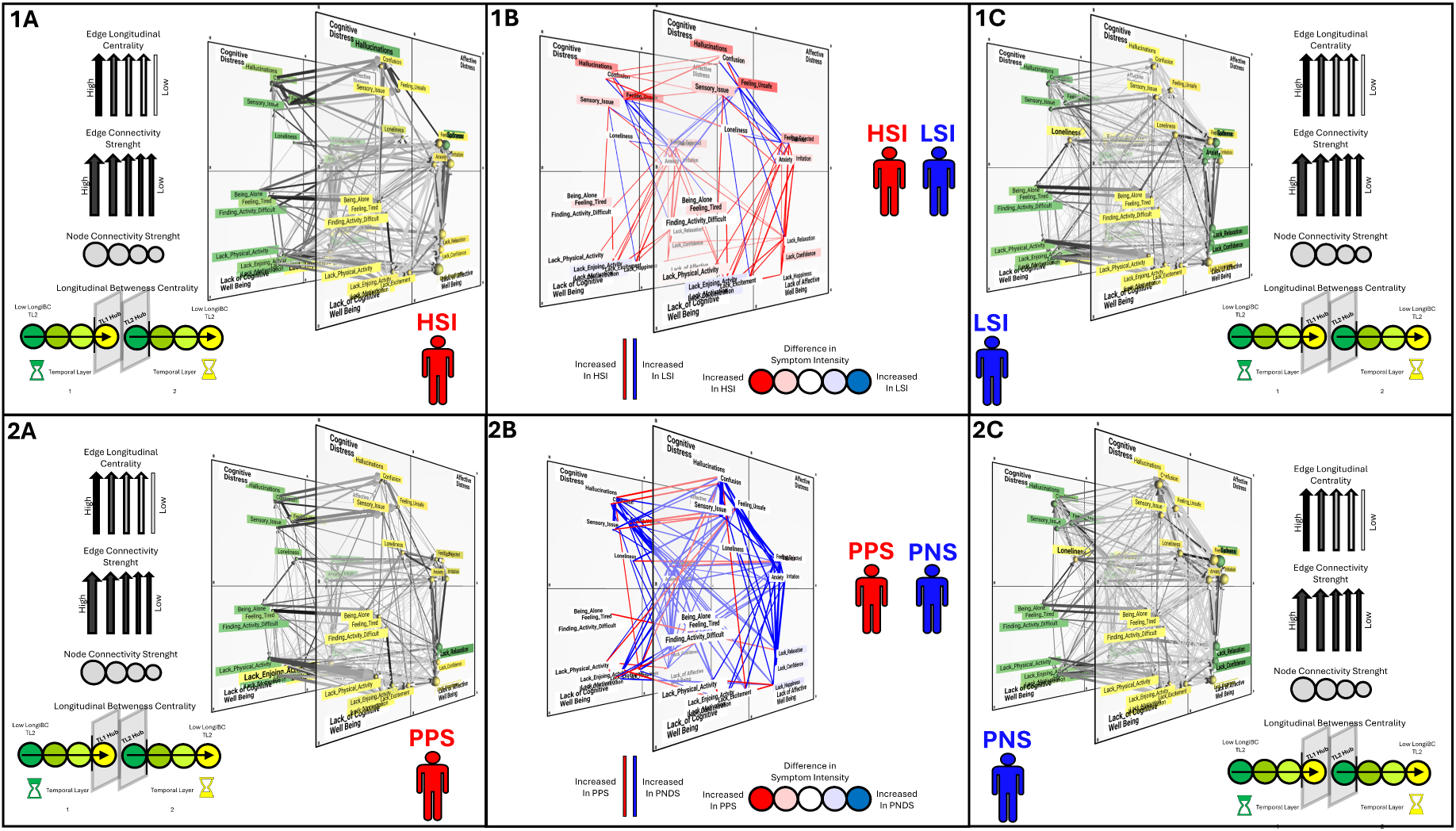
**Panels 1A-2A-1C-2C** depict Multi-Layer Temporal Networks constructed in the 4 populations separately **Panel-1A** High-Symptom-Intensity network, **Panel-1C** Low-Symptom-Intensity network, **Panel-2A:** Predominantly-Positive-Symptom network **Panel-2C:** Predominantly-Negative-Symptom network. Cross-sectional connections are represented within each temporal layer. Longitudinal connections are represented along the Z axis and bridge variables located in Temporal Layer 1 (TL-1) represented on the left to variables located in Temporal Layer 2 (TL-2) represented on the right. Node position within each temporal layer is determined according to the structure of cross-sectional networks as defined by network dimensionality reduction described elsewhere, grouping together variables that are more tightly related cross-sectionally. The slope of longitudinal edges reflects the propensity for dynamic interactions between qualitatively different psychological-contextual states occurring across temporal assessments. Thickness of network edges represents the strength of statistical associations between variables derived from cross-sectional or longitudinal mixed-model-linear regression. Arrows on connections are derived from shortest path analysis applied to Multi-Layer Temporal Network (MLTN) and indicate that a particular edge, is part of a shortest path connecting variables across temporal layers. Transparency of network edges reflects the number of shortest paths that traverse through it. Node color reflects Longitudinal-Betweenness-Centrality. Green to Yellow color-coding is reverse coded in TL-1and TL-2 so as to visually represent psychological-contextual pathways flowing from Past States represented in green to future states represented in yellow. Specifically, TL-1 Variables are shaded from green to yellow according to their propensity to act as gateways towards future states in TL-2. TL-2 variables are shaded from yellow to green according to their propensity to act as funnels from past states in TL-1. Variables that act as significant longitudinal hubs are highlighted in bold. Node size represents the summed connectivity strength of the corresponding psychological or contextual variable. **Panels 1B and 2B** depict Multi-Layer Temporal Networks constructed to illustrate network edges the strength of which is significantly influenced by SIPS-Dimensions. The network structure maintains the same longitudinal format as in Panels 1A-2A-1C-2C, with cross-sectional connections represented within each temporal layer and longitudinal connections bridging variables from Temporal Layer 1 (TL-1) on the left to Temporal Layer 2 (TL-2) on the right. Color-coding of network edges indicates the direction with which SIPS-Dimensions influence association strength between corresponding EMA variables. Color-coding of network nodes indicates the direct with which SIPS-Dimensions influence the average intensity of corresponding EMA variables. **Panel 1B:** Illustrates network edges that are modulated by SIPS-Dimension-1 capturing overall positive and negative symptom intensity. Red Edges indicate connections that increased in strength in High-Symptom-Intensity, while Blue Edges indicate connections that are stronger in Low-Symptom-Intensity. Red Nodes indicate variables that increased in intensity in High-Symptom-Intensity, while Blue Nodes indicate variables that are increased in intensity in Low-Symptom-Intensity. **Panel 1C:** Illustrates network edges that are modulated by SIPS-Dimension-2 capturing the differential effects of Predominantly-Positive-Symptoms vs Predominantly-Negative Symptoms. Red Edges indicate connections that increased in strength in Predominantly-Positive-Symptoms, while Blue Edges indicate connections that are stronger in Predominantly-Negative-Symptoms. Red Nodes indicate variables that increased in intensity in Predominantly-Positive-Symptoms, while Blue Nodes indicate variables that are increased in intensity in Predominantly-Negative-Symptoms. **Links to 3d interactive visualization:** https://dev.mlnetwork-diplab.ch/3dvisualizer/net_dim1_hsi/ https://dev.mlnetwork-diplab.ch/3dvisualizer/net_dim1_lsi/ https://dev.mlnetwork-diplab.ch/3dvisualizer/net_dim2_pns/ https://dev.mlnetwork-diplab.ch/3dvisualizer/net2_dim2_pps/ https://dev.mlnetwork-diplab.ch/3dvisualizer/dim2_pns_pps/ https://dev.mlnetwork-diplab.ch/3dvisualizer/dim1_lsi_hsi/

SIPS-Dimension-1 capturing overall symptom severity, was associated with weaker cross-sectional connections between Psychotic/Cognitive-Distress and Affective-Distress EMA items, while cross-sectional connections among Psychotic/Cognitive-Distress items (*Confusing-Reality-with-Imagination* and *Feeling-Unsafe)* were stronger. EMA Psychotic/Cognitive-Distress symptoms were furthermore more strongly predictive of each other across time and somewhat less tightly predicted by previous Affective-Distress. Affective-Distress symptoms were instead more strongly associated with both concomitant and future lack of well-being states (Figure-3-Panel-1B).

SIPS-Dimension-2, modulated several of the same alterations in EMA-network structure as SIPS-Dimension-1, capturing the relative contribution of predominantly-positive vs predominantly-negative symptoms to such alterations. In particular, SIPS-Dimension-2 modulated similar strengthening of cross-sectional and longitudinal connections between Psychotic/Cognitive-Distress items, as well as the dissociation of these items from Affective-Distress, demonstrating that such alterations were associated with the presence of Predominantly-Positive-Symptom (PPS) patterns (Figure-3-Panel-2B). This would suggest that in individuals with more severe SIPS positive psychotic symptoms, who clustered in HSI and PPS subgroups, EMA Psychotic/Cognitive-Distress experiences were more not only more intense, but also fluctuated as more coherent and temporally stable events, that were less tightly linked to other forms of psychological distress.

SIPS-Dimension-2 also modulated the strengthening of connections between Affective-Distress and Lacking-Affective-Wellbeing variables observed for SIPS-Dimension-1, demonstrating that such alterations were specifically driven by individuals with PNS. Moreover, in PNS individuals Lacking-Affective-Wellbeing variables were also more tightly predicted by previous Affective-Distress. This would suggest that severity of SIPS negative symptoms was reflected both in the average intensity of EMA psychological well-being states, and in the dynamic phenomenology of such states, which were more tightly associated with concomitant and previous Affective-Distress.

### Dissecting psychological contextual dynamics linked to positive-negative dimensions with Behavioral Tractography

We then explored the potential of a recently proposed Behavioral-Tractography approach to dissect specific behavioral pathways linked to different SIPS symptom dimensions. Behavioral-Tractography performed on the entire 22q11DS sample, revealed nine distinct bundles composed of behavioral pathways mediating qualitatively similar dynamic behavioral interactions, as reflected by similar three-dimensional trajectories. Both the composition and the average trajectory of such Behavioral-Tractography-Bundles were highly consistent with previous observations in independent typically developing and clinical samples (Supplementary Analysis 1). The average trajectory of such Behavioral-Tractography-Bundles provided a quantitative and graphical description of the main types of dynamic interactions between psychological states.

**Bundles 1-2** connected psychological distress states in the upper portion of the network across time.

**Bundle-1**: TL1-Affective-Distress → TL2-Affective-Distress, mediated by TL1-*Lacking Relaxation*, TL1-*Lacking Happiness*, TL2- *Lacking Relaxation*, TL2- *Sadness*.

**Bundle-2**: TL1-Cognitive-Distress → TL2-Affective-Distress, mediated by TL1- *Feeling Unsafe*, TL1- *Hallucinations*, TL2- *Hallucinations*

**Bundles 3-4** connected lack of psychological well-being states in the lower portion of the network across time.

**Bundle-3**: TL1-Lacking-Affective-Wellbeing → TL2-Lacking-Affective-Wellbeing, mediated by TL1-*Lacking-Happiness* and TL1-*Lacking Confidence*.

**Bundle-4**: TL1-Lacking-Affective-Wellbeing → TL2-Lacking-Cognitive-Wellbeing (LCWB), mediated by TL1-*Lacking Motivation*, TL2- *Lacking Physical Activity*, TL2- *Lacking Enjoying Activity*, TL2- *Lacking Concentration*.

**Bundles 5-7** connected lack of psychological well-being to psychological distress.

**Bundle-5:** TL1-Lacking-Affective-Wellbeing → TL2-Cognitive-Distress, mediated by TL1-*lacking Happiness*, TL1-*Feeling Rejected*, TL2-*Anxiety*, TL2-*Sadness*.

**Bundle-6:** TL1-Lacking-Cognitive-Wellbeing → TL2-Affective-Distress, mediated by TL2-*Lacking-Happiness*, TL2-*Lacking Confidence* and TL2-*Feeling Rejected*.

**Bundle 7**: TL1-Lacking-Cognitive-Wellbeing → TL2-Cognitive-Distress, mediated by TL1- Social Isolation (*Being Alone*, *Loneliness*), TL1/TL2-*Feeling Tired*, TL2-*Being Alone*

**Bundles 8-9** connected psychological distress to lack of psychological well-being.

**Bundle 8**: TL1-Cognitive-Distress → TL2-Lacking-Affective-Wellbeing, mediated by TL1- *Loneliness*, TL1- *Anxiety*, TL1-*Sadness*, TL1-*Being Alone*, TL2-*Lacking Relaxation*, TL2-*Anxiety*, TL2-*Sadness*.

**Bundle 9**: TL1-Cognitive-Distress → TL2-Lacking-Cognitive-Wellbeing mediated by TL1-*Lacking Relaxation*, TL1-*Irritation*, TL1-*Feeling Rejected*

These bundles, (described in detail in Supplementary-Figure-1), highlight the complex interplay between different psychological states over time, revealing key mediating factors in the transitions between states.

We then performed a two-population Behavioral-Tractography, to identify population-specific behavioral trajectories differentiating SIPS-dimension subgroups. We applied Behavioral-Diffusion-Analysis to pinpoint individual variables driving differences in Bundle trajectories across populations. Here we provide a brief description of population specific trajectories which is detailed in Figures-4/5, along with links to a dedicated online 3D platform facilitating exploration of results. BDA results are detailed in Supplementary-Table-1 while Figure-4/5-Panel-10A/B depicts population-specific BDA vectors of variables, driving differences in Bundle Trajectories.

**Figure 4:**
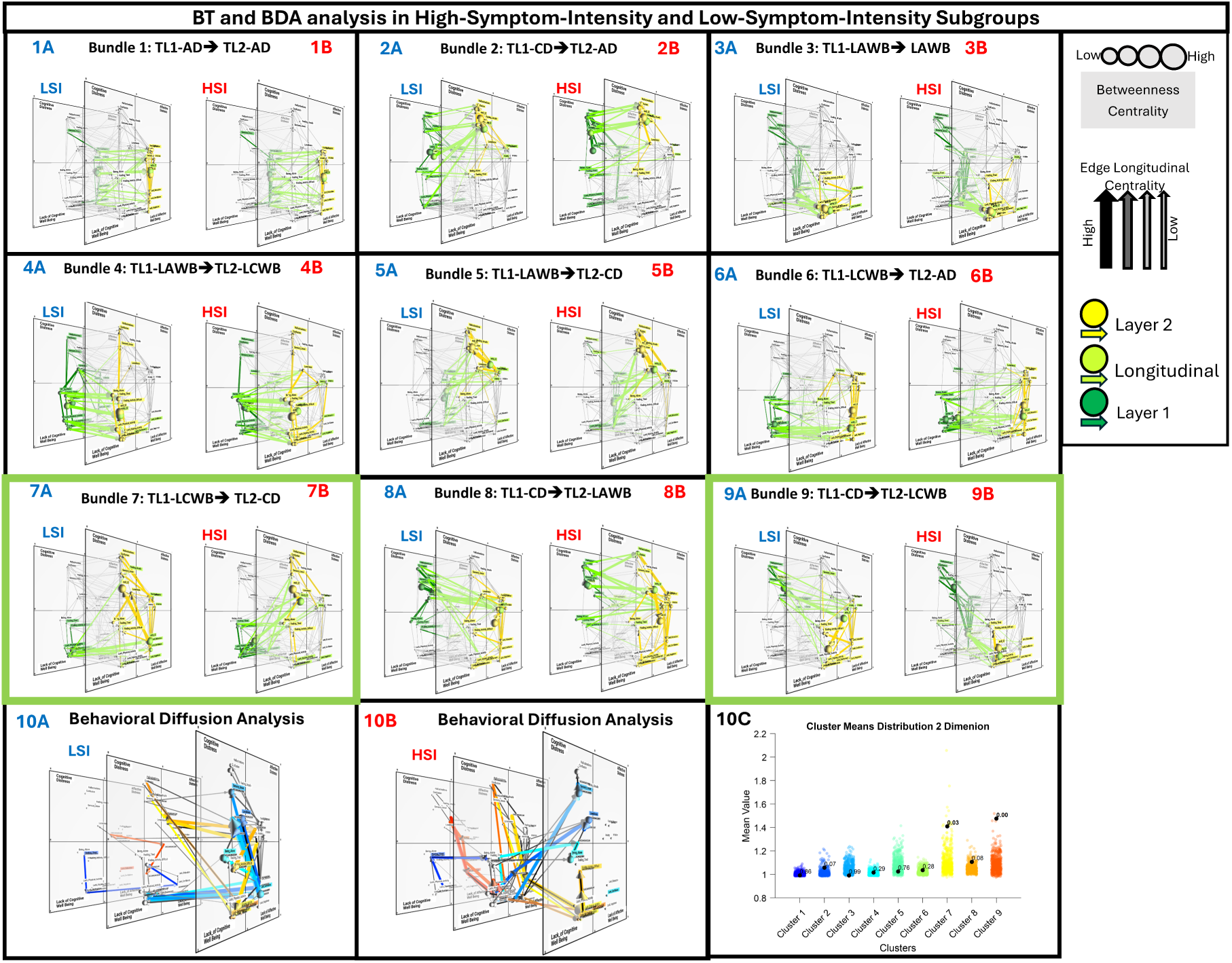
Comparison of population-specific trajectories of Behavioral Tractography (BT) Bundles in High-Symptom-Intensity bs Low-Symptom-Intensity subgroups defined according to SIPSDimension-1. **Panels 1-9:** Results of Behavioral-Tractography trajectory for each of 9 Bundles in High-Symptom-Intensity (A)and Low-Symptom-Intensity(B) subgroups; Green to Yellow color coding reflects the dynamic progressing of paths from TL1 to TL2 (Start-TL1 → Exit-TL1: Dark Green; Exit-TL1 → Entry-TL2: Light Green; Entry-TL2 → End-TL2: Yellow). Panels 1A-1B: Bundle 1: TL1-AD→TL2-AD. Panels 2A-2B: Bundle 2: TL1-CD→TL2-AD. Panels 3A-3B: Bundle 3: TL1-LAWB→TL2- LAWB. Panels 4A-4B: Bundle 4: TL1- LAWB→TL2-LCWB. Panels 5A-5B: Bundle 5: TL1- LAWB →TL2-CD. Panels 6A-6B: Bundle 6: TL1-LCWB→TL2-AD. Panels 7A-7B: Bundle 7: TL1- LCWB →TL2-CD. Panels 8A-8B: Bundle 8: TL1-CD→TL2-LAWB. Panels 9A-9B: Bundle 9: TL1-CD→TL2-LCWB. **Panels 10A-10B**: Results of BDA for variables in LSI (Panels 10A) and HSI (Panels 10B). Warm colors identify variables that drive significant BT differences in Bundle-9 (TL1-LCWB→TL2-CD). Cold colors identify variables that drive significant BT differences in Bundle-7 (TL1-CD→TL2-LCWB). BDA trajectories reflecting differential functional role of TL1-Variables connect TL1 to TL2, while BDA trajectories reflecting differential functional role of TL2-Variables connected TL2 to TL3. The individual PC paths that contribute to average BDA trajectories are represented in Black. **Panel 10C:** Statistical comparison of differences in the average trajectory of each of 9 Behavioral-Tractography Bundles across High-Symptom-Intensity and Low-Symptom-Intensity Populations. For each bundle (represented by a column), we calculated the ratio between the average Euclidian distance between Bundle-specific pathways across populations, capturing how dissimilar the trajectory of a given Bundle is across populations, and the average Euclidian distance separating Bundle-specific pathways in each population, capturing within-population heterogeneity of the BT-Bundle. To assess the statistical significance of these differences, we performed a permutation test. The colored dots in each column depict the null distribution generated from 500 permutations, where subjects were randomly reassigned between groups. The p-value adjacent to each black dot indicates the proportion of permutations yielding a ratio equal to or greater than the observed value. Bundles with lower p-values (e.g., p < 0.05) represent path structures that significantly differentiate between populations while maintaining within-population consistency, potentially reflecting distinct symptom-interaction dynamics between the groups. **Links to 3d interactive visualization**: **HSI:** https://dev.mlnetwork-diplab.ch/3dvisualizer/dim1_dim1_cl1/ https://dev.mlnetwork-diplab.ch/3dvisualizer/dim1_dim1_cl2/ https://dev.mlnetwork-diplab.ch/3dvisualizer/dim1_dim1_cl3/ https://dev.mlnetwork-diplab.ch/3dvisualizer/dim1_dim1_cl4/ https://dev.mlnetwork-diplab.ch/3dvisualizer/dim1_dim1_cl5/ https://dev.mlnetwork-diplab.ch/3dvisualizer/dim1_dim1_cl6/ https://dev.mlnetwork-diplab.ch/3dvisualizer/dim1_dim1_cl7/ https://dev.mlnetwork-diplab.ch/3dvisualizer/dim1_dim1_cl8/ https://dev.mlnetwork-diplab.ch/3dvisualizer/dim1_dim1_cl9/ https://dev.mlnetwork-diplab.ch/3dvisualizer/qpa_dim1_hsi/ **LSI:** https://dev.mlnetwork-diplab.ch/3dvisualizer/dim1_dim2_cl1/ https://dev.mlnetwork-diplab.ch/3dvisualizer/dim1_dim2_cl2/ https://dev.mlnetwork-diplab.ch/3dvisualizer/dim1_dim2_cl3/ https://dev.mlnetwork-diplab.ch/3dvisualizer/dim1_dim2_cl4/ https://dev.mlnetwork-diplab.ch/3dvisualizer/dim1_dim2_cl5/ https://dev.mlnetwork-diplab.ch/3dvisualizer/dim1_dim2_cl6/ https://dev.mlnetwork-diplab.ch/3dvisualizer/dim1_dim2_cl7/ https://dev.mlnetwork-diplab.ch/3dvisualizer/dim1_dim2_cl8/ https://dev.mlnetwork-diplab.ch/3dvisualizer/dim1_dim2_cl9/ https://dev.mlnetwork-diplab.ch/3dvisualizer/qpa_dim1_lsi/

Both SIPS-Dimension-1 (p=0.03) and SIPS-Dimension-2 (p<0.001) modulated the trajectory of Bundle-7 connecting TL1-Lacking-Cognitive-Wellbeing to TL2-Psychotic/Cognitive-Distress, defining 4 population specific trajectories.

**SIPS-Dimension-1-LSI** (Figure-4-Panel-7A): Effects of TL1-Lacking-Cognitive-Wellbeing to TL-2-Psychotic/Cognitive-Distress indirectly mediated by TL2 Lacking-Affective-Wellbeing (TL2-*Lacking-Confidence*).

**SIPS-Dimension-1-HSI** (Figure-4-Panel-7B): Direct pathway linking TL1-Lacking-Cognitive-Wellbeing (*TL1-Feeling-Tired*) to TL2 social isolation (TL2-*Loneliness*, TL2-*Being-Alone*) and TL2-Psychotic/Cognitive-Distress (TL2-*Sensory-Issue*).

**SIPS-Dimension-2-PNS** (Figure-5-Panel-7A): TL1-Social-Isolation predicts TL2-Psychotic/Cognitive-Distress (TL2-*Feeling-Unsafe*, TL2-*Hallucinations*, TL2-*Confusing-Reality-with-Imagination)*.

**Figure 5:**
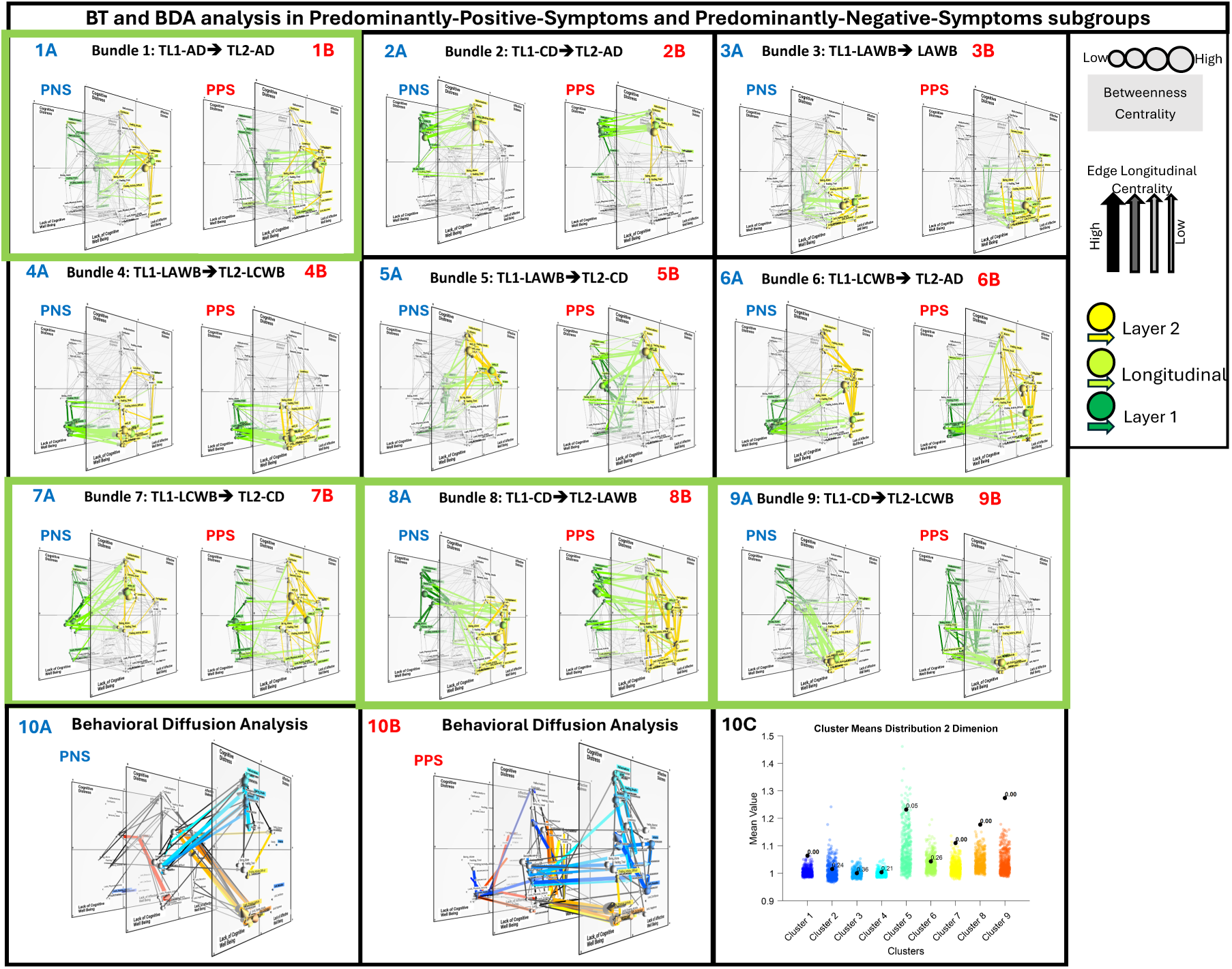
**Panels 1-9:** Results of Behavioral-Tractography trajectory for each of 9 Bundles in Predominantly-Positive-Symptoms (A) and Predominantly-Negative-Symptoms(B) subgroups; Green to Yellow color coding reflects the dynamic progressing of paths from TL1 to TL2 (Start-TL1 → Exit-TL1: Dark Green; Exit-TL1 → Entry-TL2: Light Green; Entry-TL2 → End-TL2: Yellow). Panels 1A-1B: Bundle 1: TL1-AD-->TL2-AD. Panels 2A-2B: Bundle 2: TL1-CD→TL2-AD. Panels 3A-3B: Bundle 3: TL1-LAWB→TL2- LAWB. Panels 4A-4B: Bundle 4: TL1- LAWB→TL2-LCWB. Panels 5A-5B: Bundle 5: TL1- LAWB →TL2-CD. Panels 6A-6B: Bundle 6: TL1-LCWB→TL2-AD. Panels 7A-7B: Bundle 7: TL1- LCWB →TL2-CD. Panels 8A-8B: Bundle 8: TL1-CD→TL2-LAWB. Panels 9A-9B: Bundle 9: TL1-CD→TL2-LCWB. **Panels 10A-10B**: Results of BDA for variables in PPS (Panels 10A) and PNS (Panels 10B). Warm colors identify variables that drive significant BT differences in Bundle-9 (TL1-LCWB→TL2-CD). Cold colors identify variables that drive significant BT differences in Bundle-7 (TL1-CD→TL2-LCWB). BDA trajectories reflecting differential functional role of TL1-Variables connect TL1 to TL2, while BDA trajectories reflecting differential functional role of TL2-Variables connected TL2 to TL3. The individual PC paths that contribute to average BDA trajectories are represented in Black. **Panel 10C:** Statistical comparison of differences in the average trajectory of each of 9 Behavioral-Tractography Bundles across Predominantly-Positive-Symptoms and Predominantly-Negative-Symptoms subgroups. For each bundle (represented by a column), we calculated the ratio between the average Euclidian distance between Bundle-specific pathways across populations, capturing how dissimilar the trajectory of a given Bundle is across populations, and the average Euclidian distance separating Bundle-specific pathways in each population, capturing within-population heterogeneity of the Behavioral-Tractography -Bundle. To assess the statistical significance of these differences, we performed a permutation test. The colored dots in each column depict the null distribution generated from 500 permutations, where subjects were randomly reassigned between groups. The p-value adjacent to each black dot indicates the proportion of permutations yielding a ratio equal to or greater than the observed value. Bundles with lower p-values (e.g., p < 0.05) represent path structures that significantly differentiate between populations while maintaining withinpopulation consistency, potentially reflecting distinct symptom-interaction dynamics between the groups. **Links to 3d interactive visualization:** **PNS:** https://dev.mlnetwork-diplab.ch/3dvisualizer/dim2_dim1_cl1/ https://dev.mlnetwork-diplab.ch/3dvisualizer/dim2_dim1_cl2/ https://dev.mlnetwork-diplab.ch/3dvisualizer/dim2_dim1_cl3/ https://dev.mlnetwork-diplab.ch/3dvisualizer/dim2_dim1_cl4/ https://dev.mlnetwork-diplab.ch/3dvisualizer/dim2_dim1_cl5/ https://dev.mlnetwork-diplab.ch/3dvisualizer/dim2_dim1_cl6/ https://dev.mlnetwork-diplab.ch/3dvisualizer/dim2_dim1_cl7/ https://dev.mlnetwork-diplab.ch/3dvisualizer/dim2_dim1_cl8/ https://dev.mlnetwork-diplab.ch/3dvisualizer/dim2_dim1_cl9/ https://dev.mlnetwork-diplab.ch/3dvisualizer/qpa_dim2_pns/ **PPS:** https://dev.mlnetwork-diplab.ch/3dvisualizer/dim2_dim2_cl1/ https://dev.mlnetwork-diplab.ch/3dvisualizer/dim2_dim2_cl2/ https://dev.mlnetwork-diplab.ch/3dvisualizer/dim2_dim2_cl3/ https://dev.mlnetwork-diplab.ch/3dvisualizer/dim2_dim2_cl4/ https://dev.mlnetwork-diplab.ch/3dvisualizer/dim2_dim2_cl5/ https://dev.mlnetwork-diplab.ch/3dvisualizer/dim2_dim2_cl6/ https://dev.mlnetwork-diplab.ch/3dvisualizer/dim2_dim2_cl7/ https://dev.mlnetwork-diplab.ch/3dvisualizer/dim2_dim2_cl8/ https://dev.mlnetwork-diplab.ch/3dvisualizer/dim2_dim2_cl9/ https://dev.mlnetwork-diplab.ch/3dvisualizer/qpa_dim2_pps/

**SIPS-Dimension-2-PPS** (Figure-5-Panel-7B): TL1-Lacking-Cognitive-Wellbeing (TL1- *Lacking-Excitement*, TL1-*Lacking-Enjoyment*), mediates the effects of TL1-Social Isolation on subsequent TL-2 Affective variables (TL2-*Lacking-Relaxation*, TL2-*Irritation)* and TL2- Psychotic/Cognitive-Distress.

Taken together, HSI and PPS Bundle-7 trajectories, would suggest that in individuals with more severe SIPS positive symptoms, Lacking-Cognitive-Wellbeing variables exert a more direct effect on subsequent EMA Psychotic/Cognitive-Distress variables, compared to a higher contribution of previous social isolation in PNS subgroup, and subsequent affective variables in LSI. Interestingly, the differential contribution of Lacking-Cognitive-Wellbeing variables was observed despite the lower intensity with which these states were reported in both the HSI and PPS subgroups, potentially indicating heighted vulnerability to develop Psychotic/Cognitive-Distress following insufficient Cognitive-Wellbeing.

Both SIPS-Dimension-1 (p<0.001) and SIPS-Dimension-2 (p<0.001) also modulated the trajectory of Bundle-9 linking TL1-Psychotic/Cognitive-Distress to TL2-Lacking-Cognitive-Wellbeing, defining 4 population-specific trajectories.

**SIPS-Dimension-1-LSI** (Figure-4-Panel-9A): Effects of TL1-Psychotic/Cognitive-Distress on TL2-Lacking-Cognitive-Wellbeing (TL2-Lacking-Enjoyment, TL2-Finding-Activity-Difficult, TL2-Lacking-Concentration, TL2-Lacking-Excitement, TL2-Lacking-Motivation) are mediated by TL1-Affective-Distress (TL1-Anxiety) and TL2-Lacking-Affective-Wellbeing.

**SIPS-Dimension-1-HSI** (Figure-4-Panel-9B): Effects of TL1-Psychotic/Cognitive-Distress on TL2-Lacking-Cognitive-Wellbeing are mediated by TL1-Lacking-Affective-Wellbeing (TL1-Lacking-Relaxation)

**SIPS-Dimension-2-PNS** (Figure-5-Panel-9A): Effects of TL1-Psychotic/Cognitive-Distress on TL2-Lacking-Cognitive-Wellbeing (TL2-Lacking-Enjoyment, TL2-Finding-Activity-Difficult, TL2-Lacking-Concentration, TL2-Lacking-Excitement, TL2-Lacking-Motivation) are mediated by TL1-Affective-Distress variables (TL1-Sadness).

**SIPS-Dimension-2-PPS** (Figure-5-Panel-9B): Effects of TL1-Psychotic/Cognitive-Distress on TL2-Lacking-Cognitive-Wellbeing are mediated by TL1-Lacking-Affective-Wellbeing (TL1-Lacking-Relaxation).

Taken together, the similar Bundle-9 trajectories in HSI and PPS subgroups, would suggest that individuals with more severe SIPS positive symptoms experience more direct transitions from previous TL1-Psychotic/Cognitive-Distress to subsequent TL2-Lacking-Cognitive-Wellbeing, occurring independently of affective distress variables.

PNS individuals instead presented a unique pathway linking TL1-Sadness to TL2-Lacking-Cognitive-Wellbeing. A similar pattern contributed to the differential PNS trajectory of BT-Bundle-8 (Figure-5-Panel-8A), where TL1-*Sadness* (p<0.001) mediated the transition from TL1-Psychotic/Cognitive-Distress and TL2-Lacking-Affective-Wellbeing. This would suggest that individuals with PNS, affective-Distress variables, in particular *Sadness*, play a more direct role in the severe reductions of psychological well-being reported by this population. Interestingly, this was observed despite the reduced intensity of *Sadness* reported in the PNS subgroup, suggesting that PNS individuals may present heightened vulnerability to severe psychological wellbeing reductions in response to relatively minor affective distress experiences. Overall, these results would suggest that the average intensity of psychological experiences may provide an incomplete, or even misleading representation of their importance in influencing fluctuations in future psychological states.

## Discussion

In the present paper, we explore the potential of a recently proposed Behavioral-Tractography approach for translating the rich dynamic and ecologically grounded view of psychological phenomena provided by EMA [15], to a deeper and clinically meaningful understanding of the pathophysiological interplay between different psychosis symptom dimensions [44].

We firstly evaluated weather EMA questionnaires could effectively capture the same multidimensional clinical phenomena characterized with gold-standard psychosis clinical interviews in 22q11DS. Indeed, such validation is a critical perquisite for drawing conclusions on psychosis symptom dynamics from EMA studies [14]. The structure of across subject variation in SIPS symptom intensity was highly consistent with previous reports [42, 43]. SIPS-Dimension-1 reflected overall severity of both positive and negative symptoms, while SIPS-Dimension-2 highlighted the differential expression of predominantly-positive vs predominantly-negative symptom patterns. The average intensity of most EMA items significantly differed across such SIPS-Dimensions. Individuals with High-Symptom-Intensity (HSI) and Predominantly-Positive-Symptom (PPS) both reported higher EMA items reflecting positive psychotic symptoms (e.g., *Hallucinations*, *Feeling-Unsafe, Confusing-Reality-with-Imagination*), while individuals with Predominantly-Negative-Symptoms (PNS) reported reduced psychological well-being—both affective (e.g., *Lacking-Happiness Lacking-Relaxation*) and cognitive/motivational (e.g., *Lacking-Motivation, Lacking-Enjoyment*). These results show that EMA questionnaires can effectively capture across-subject variation in both positive and negative psychosis symptomatic dimensions, defined with gold standard clinical interviews.

To further evaluate EMA questionnaire validity, we explored whether traditional psychosis dimensions would map on to the low-dimensional structure of dynamic psychological phenomena, characterized by EMA Network-Dimensionality-Reduction (NDR) [32]. NDR results were highly consistent with previous literature[32]. The correlation structure of behavioral fluctuations was organized along two rationality and valence dimensions, defining four Affective-Distress, Cognitive-Distress, Lacking-Affective-Wellbeing and Lacking-Cognitive-Wellbeing network quadrants. Remarkably, the ways in which EMA items differed according to SIPS dimensions, was mirrored by their differential mapping along such NDR dimensions. EMA-correlates of positive symptoms clustered in the upper-left Cognitive-Distress quadrant, while correlates of negative symptom clustered in distinct lower-left Lacking-Cognitive-Wellbeing and lower-right Lacking-Affective-Wellbeing quadrants. The alignment between SIPS-EMA correlations and NDR dimensions, which can be defined as trait-state homomorphy[45], suggests that latent factors underlying inter-individual variability of psychotic symptoms severity, may be tied to mechanisms underpinning dynamic fluctuations in symptom severity experienced by individual patients across time. This novel observation could carry significant implications for trait-vs-state models of psychosis pathophysiology [46, 47]. Specifically, it would suggest that psychosis symptoms intensity characterized by traditional clinical assessments, does not measure a static level of disease severity, but rather reflects the aggregate of inherently dynamic fluctuations in different symptom dimensions [46, 47]. Our results suggest that EMA questionnaires can effectively capture dynamic fluctuations in the same phenomena measured with current gold-standard static psychosis assessments [14].

The second objective of the present study was to explore EMA’s potential in generating additional clinically relevant insights on behavioral dynamics associated with different psychosis symptom dimensions. We employed an innovative 3D-Temporal-Multilayer-Network approach, providing an integrated representation of cross-sectional co-occurrence and dynamic symptom interactions unfolding across time, which revealed distinct behavioral dynamics associated with SIPS positive symptom severity. Specifically, both HSI and PPS subgroups, presented stronger correlations between EMA Psychotic/Cognitive-Distress items, while associations between psychotic and affective distress variables were weaker. This suggests that in more severely affected individuals, psychotic experiences (e.g., hallucinations, paranoia, reality confusion), are not only more intense, but also manifest as more multifaceted symptom constellations, that are increasingly dissociated from other forms of psychological distress. The increasing dissociation of cognitive and affective distress variables with increasing psychosis severity, supports the value of the cognitive-to-affective *rationality* NDR dimensions in differentiating psychosis from other forms of psychological distress. Indeed, mapping of psychotic symptoms as cognitive distress, strikingly recapitulates phenomenological descriptions of the psychotic symptoms as a “cognitive” attempt to make sense of internally or externally generated events that are experienced as unusual, bizarre, or unexpected [48].

The 3D-Temporal-Multilayer-Network approach further reveled that positive symptom severity exerted a combined impact on both cross-sectional correlations, and dynamic symptom interactions occurring from one assessment to the next. Indeed, HSI and PPS subgroups presented strengthening of longitudinal connections linking psychotic symptoms across time, suggesting that in more symptomatic subjects, psychotic symptoms become increasingly self-sustaining and independent from affective or contextual fluctuations. Interestingly, these results are highly consistent with Bayesian models of psychosis, according to which reality distortion underlying positive symptoms, may stem from as an inability to flexibly adapt internal models of the world, based on dynamically fluctuating experience of recent contextual factors [49]. Of note, according to Bayesian models individuals with ASD may present an opposite tendency to rely more strongly on recent sensory information [50–52], which would prove advantageous in repetitive environments requiring high sensory precision [53], but may also contribute to higher psychological volatility in response minor variation in contextual factors [52]. In accordance with such models, we recently demonstrated that ASD and 22q11DS populations present opposite network alterations, compared to typically developing individuals, reflecting diametrical differences in psychological-contextual dynamics. Our current results further support such Bayesian models, by demonstrating that differences in psychological-contextual dynamics observed across 22q11DS and ASD populations accentuate as psychosis severity increases within the 22q11DS population. Overall, these results suggest that measuring the short-term dynamics of psychosis phenomenology, can provide additional insights on both the subjective experience and underlying pathophysiology of positive psychotic symptoms.

The unique comprehensive clinical assessment and analysis pipeline revealed dynamic behavioral alterations, which were not limited to positive symptoms, and specifically reflected SIPS negative symptom severity. Indeed, both HSI and PNS subgroups, presented stronger connections between EMA-correlates of negative symptoms (Lacking-Affective-Wellbeing and Lacking-Cognitive-Wellbeing variables in lower-left and lower-right quadrants), and Affective-Distress states (*Sadness, Irritation, Anxiety,* upper-right quadrant). This would suggest that in individuals with more severe negative symptoms, reductions of psychological well-being are not only more severe, but also more tightly related to concomitant affective distress. HSI and PNS subgroups also presented a widespread strengthening of time-lagged connections, which primarily affected “diagonal” connections linking different symptomatic dimensions across temporal layers. This would suggest that negative symptom severity may be linked to increased temporal dependencies between past and future psychological states, which however did not reflect the temporal inertia of more inherently stable states, but rather stemmed from an increased propensity to dynamically shift from one symptomatic dimension to another across time.

We employed the Behavioral Tractography pipeline to shed on such population-specific pathways underlying dynamic shifts between different symptom dimensions[32]. Interestingly, both SIPS Dimensions influenced bi-directional interactions between Cognitive-Distress positive symptoms, and Lacking-Cognitive-Wellbeing negative symptoms, as captured by population-specific trajectories of Behavioral-Bundles 7 and 9. The HSI subgroup, characterized by combined increase of positive and negative symptoms, presented more direct bidirectional trajectories between such states, while in the LSI subgroup such interactions were mediated by affective variables. This would suggest the combined presentation of positive and negative symptoms, may stem from a tighter dynamic interaction between states, rather than from co-occurrence of independent disease mechanisms. Of note, this tighter relationship was not reflected in the strength of cross-sectional correlations, suggesting that individuals with combined positive and negative symptomatology, still experience these symptoms as phenomenologically distinct events, occurring at different moments in time. Instead, the HSI subgroup presented heighted propensity to develop *secondary* psychotic experiences following previous motivational deficits, as well as secondary motivational deficits following previous psychotic cognitive distress.

The fine-grained Behavioral-Tractography pipeline further revealed a differential modulation of dynamic pathways linking EMA positive and negative symptoms, exerted by SIPS Predominantly-Positive vs Predominantly-Negative symptom patterns. In the PNS subgroup, the interaction between previous Lacking-Cognitive-Wellbeing and future Cognitive-Distress, described by Bundle-7, was more strongly mediated by social isolation factors, compared to a more direct effect of Lacking-Cognitive-Wellbeing, observed in PPS individuals. As such, in individuals with more severe positive symptoms, fluctuations in psychotic symptom severity may be more independent of contextual factors, in accordance with previously highlighted Bayesian models [49]. The PNS subgroup was instead characterized by direct pathways linking, previous feelings of sadness associated with psychotic/cognitive-distress to subsequent reductions in affective and cognitive well-being states, described by Behavioral-Bundles 8 and 9. These results would suggest that increased negative symptom severity, may stem from a heightened propensity to develop secondary reductions in motivation and affective well-being, following previous affective distress. Of note, these findings would challenge the view of negative symptoms as stable expressions of disease progression, that become increasingly intractable as their severity increases [5, 54, 55]. Indeed, not only did negative symptom intensity fluctuate at short-time frames, but such symptomatic dynamics were accentuated in more severely affected individuals, due to stronger modulation exerted by previous affective distress experiences.

To our knowledge this is the first study to comprehensively investigate dynamic short-term interactions between the different symptom dimensions of psychosis. Our results are however consistent with a growing body of literature demonstrating that both positive and negative symptom dimensions can fluctuate dynamically in relation to external contextual factors and other psychological states [20, 22, 56–59]. Specifically, our results align with recent EMA studies[20, 22, 57, 58], suggesting that positive and negative symptom can interact at much faster temporal dynamics than was traditionally conceptualized [41], which could carry important pathophysiological implications. Indeed, current neurobiological models struggle to reconcile the proposedly opposite hyper-dopaminergic underpinnings of positive symptoms, with the dopaminergic hypoactivity thought to contribute to motivational deficit dimension of negative symptoms[10]. Our results suggest that individuals presenting with combined positive symptoms and motivational deficits, do not actually experience these symptoms concomitantly, but rather sequentially, through short-term temporal interactions. This could suggest that their opposite hyper/hypo dopaminergic underpinnings might potentially emerge from homeostatic regulation of striatal dopaminergic signaling [60–62] that has been shown to play a key role in rapidly fluctuating secondary motivational deficits [63].

At a broader level, our findings suggest that vulnerability to downstream psychological consequences may vary independently of the average intensity with which symptoms are experienced. Indeed, HSI and PPS individuals presented increased vulnerability to develop secondary positive psychotic symptoms following previous motivational deficits, despite reporting on average milder motivational symptoms. Similarly, in PNS individuals, vulnerability to develop secondary negative symptoms following previous affective distress was observed despite reduced intensity with which sadness was reported, suggesting heightened predisposition to develop psychological consequences of affective distress, rather than an increased exposure to such experiences. This would suggest that measuring dynamic behavioral interactions, could be critical to dissect mechanisms underpinning, not only severity, but also differential vulnerability to psychological experiences across subjects[64]. From a clinical perspective, assessing behavioral dynamics could be essential to capture the relative importance of psychological variables in influencing fluctuations in future psychological states [65].

From a clinical perspective, these findings would hence support the added value of EMA approaches for improving assessment and management of psychosis [16]. However, increasing the precision and detail of psychiatric assessments carries a concrete risk of drowning clinicians in information that is ultimately too complex to reliably inform clinical reasoning [66, 67]. It has been suggested that in current practice clinicians confront such complexity by flexibly combining a large-scale analysis of broad clinical patterns, similar to identifying the outline of the forest, with a second higher-level analysis of the role of individual clinical or contextual factors [68]. The Behavioral-Tractography pipeline is specifically designed to accommodate this process which could potentially contribute to increase interpretability and clinical value of EMA. NDR dimensions provide a novel large-scale mapping of dynamic psychological fluctuations, that was both sufficiently conserved to allow intuitive comparisons across clinical populations, and sufficiently detailed to differentiate psychosis symptom dimensions within the 22q11DS sample. Behavioral-Tractography builds upon this, to derive a unique multiscale-scale view of both the main types of dynamic behavioral interactions unfolding in daily life and of the role individual symptoms. In a previous study, we demonstrate that Behavioral-Tractography can differentiate 22q11DS individuals, with a known ground-truth genetic differences, from controls and individuals with idiopathic ASD [32]. Here we demonstrate the potential of Behavioral-Tractography to provide additional insight on psychological contextual dynamics underpinning different symptom dimensions within the 22q11DS population. These results support the value of Behavioral-Tractography for elucidating differential pathophysiology of conditions that might otherwise appear homogenous based on current retrospective psychiatric assessments, which could potentially contribute to improved clinical management [64, 69].

## Methods

### Sample

68 22q11DS carriers (31% female, mean age = 19.34, SD = 5.18) were included in this study and recruited through the 22q11DS Swiss longitudinal cohort [70, 71]. The majority (84% of subjects) overlapped with the 22q11DS sample described in the previously cited Behavioral-Tractography study, while 19% of participants of the previous study who lacked full clinical assessment were excluded here [32]. All participants had a confirmed genetic diagnosis of microdeletion 22q11.2 (determined by fluorescence in situ hybridization, multiplex ligation-dependent probe amplification, or micro-array analysis). Demographic details are described in Table 1.

The presence of DSM-5 psychiatric disorders was assessed with the Diagnostic Interview for Children and Adolescents-Revised [72] or Schedule for Affective Disorders and Schizophrenia for School-Age Children Present and Lifetime Version [73] (K-SADS-PL DSM-5) for participants under 18 years old and Structured Clinical Interview for DSM-IV Axis I (SCID-I) [74] or DSM-V (SCID-5-CV) [75] for participants above 18 years old.

The presence and severity of positive, negative, disorganization, and general symptoms of psychosis were evaluated along 7-point Likert scale (0-6), using the Structured Interview for Psychosis-Risk Syndromes [76]. We considered only items that were reported with non-zero intensity in > 10% of patients, which resulted in excluding the Grandiosity (P3) item of the SIPS. This aligns with established literature [14] which suggests that grandiosity is rarely reported by patients with 22q11 deletion syndrome.

### Ecological Momentary Assessment

Ecological momentary assessment (EMA) was performed with the RealLifeExp smartphone application [77]. The protocol, which was described in previous publications [13], consisted of semi-random notifications delivered eight times daily between 7:30 AM and 10:00 PM over a six-day period (maximum of 48 prompts). A minimum interval of 30 minutes was scheduled between consecutive notifications. Participants were granted a maximum of 15 minutes to initiate the questionnaire and unlimited time for completion. In the current study, in total 20 items, evaluated on a 7-point Likert scale were used.

The EMA questionnaire provided a comprehensive assessment of participants’ psychological states. It assessed psychological distress (*Sadness, Anxiety, Anger*) and wellbeing (*Happiness, Confidence, Excitement, Relaxation, Motivation*), with wellbeing items reverse-coded for analysis. It also assessed the presence of psychotic-like experiences (*Hallucinations, Confusing-Reality-with-Imagination, Feeling-Unsafe*). Additionally, participants reported contextual details such as social setting (*Being-Alone* vs with others) and activity (current task). They also rated their experiences of social context (*Feeling-Lonely, Feeling-Rejected)* and activity (*Enjoyment, Concentration, Struggling*). See Supplementary Material for the full EMA questionnaire.

## Data analysis

### Correlation between SIPS clinical patterns and EMA item intensity

We started by characterizing the correlation between clinical patterns measured with the gold-standard SIPS clinical interview and the intensity of psychological states measured in everyday life with EMA.

Previous EMA validation studies have correlated specific EMA items with the severity of the corresponding symptoms measured with traditional clinical interviews [77]. For instance, the intensity of EMA-estimated hallucinations has shown to correlate with SIPS gold-standard clinical assessment of hallucination severity [77].

Here, we aimed to provide a more comprehensive data-driven view of the correspondence between multidimensional clinical patterns measured with SIPS and psychological phenomena measured with EMA. To this end, we firstly applied Principal Component Analysis (PCA) implemented with the MATLAB (MATLAB Version: R2024a) *pca* function, to the SIPS data. We retained PCA dimensions that explained a significant portion (>10%) of the total variance, capturing coherent clinical patterns across subjects. We derived SIPS-Dimension scores capturing the extent to which the corresponding SIPS clinical pattern was expressed in each participant, which we correlated with the average intensity of each EMA item. We also applied k-means clustering using the MATLAB *k-means* function to identify subgroups that differentially expressed such SIPS-Dimensions and compared both SIPS and EMA item intensity across the resulting subgroups using a two-sample t-test.

### Characterization of EMA Behavioral dynamics through Behavioral Tractography

We then investigated whether differences in SIPS clinical patterns would also be associated with differences in patterns of dynamic interaction between behavioral states measured with EMA.

To this end, we employed a recently developed Behavioral Tractography framework which provides a quantitative and intuitive characterization of behavioral dynamics [32]. The BT framework is described in detail in a separate publication and is an open-source toolbox https://github.com/andreaimparato/Behavioral-Tractography-Toolbox, while a brief overview of the main steps of the analysis pipeline is provided below.

We first estimated the strength of associations between pairs of EMA items as the slope coefficient of a Linear Mixed-Effects Regression (MMLR) including a random intercept to account for across-subject variation in item intensity.

We estimated association strength between cross-sectional variables from the same EMA assessment as the slope coefficient of a Linear Mixed Regression (LMR) with a random intercept for each subject; formalized as follows: *Y _ij_*=*β*_0_ + *β*_1_ *X _ij_* +*u_j_* +*ε_ij_* where *Y _ij_* is the outcome variable for subject *j* at observation *i, X_ij_* is the predictor variable for subject *j* at observation *i, β*_0_ is the fixed intercept, *β*_1_ is the fixed slope, *u _j_* is the random intercept, *ε_ij_* is the residual error term, using the *lme* function in MATLAB. By computing MMLR for each pair of EMA items we constructed a 20x20 adjacency matrix. Associations that were not significant at p<0.05 after Benjamini Hochberg multiple comparisons correction [78] were set to 0. This matrix can be thought of as a network in which nodes are EMA items, and the edges capture the association strengths between nodes.

Next, we applied a Network Dimensionality Reduction (NDR) technique [29] based on PCA, implemented through MATLAB *pca* function, to the above-mentioned adjacency matrix, in order to identify large-scale dimensions capturing the differential propensity for EMA items to manifest together. The respective network nodes for each of the EMA items were then positioned in a two-dimensional space according to their loading along the two main NDR dimensions. We estimated the accuracy of such 2-dimensinal mapping as the negative correlation between the Euclidian distance separating network nodes and the association strength between corresponding EMA items.

We then built upon such 2-dimensional mapping of cross-sectional associations to develop a 3-dimensional view of the dynamics of interactions between EMA items unfolding across time. Analogous to the cross-sectional associations, we measured the strength of associations between EMA items across consecutive assessments using Time-lagged MMLR models. We then integrated these lagged associations with the previously identified cross-sectional relationships, yielding a comprehensive 40x40 multi-layer adjacency matrix that captured both current and lagged associations between variables which could then be graphically represented as a 3-dimensional multi-layer network (3D-MLN). Cross-sectional connections were represented in separate temporal layers in which nodes/symptoms were positioned along X/Y axes according to their differential propensity to manifest together, as defined by NDR. Temporal Layers were separated along a temporal Z axis which was populated by longitudinal time-lagged edges connecting EMA items from one temporal assessment to the next. These 3D-MLN were represented with a dedicated open-source network (mlnetwork-diplab.ch) visualization software that is integrated with the BT analysis toolbox. For the main figures of the present study, we included a link to the software allowing interactive 3D manipulation of networks, hence facilitating in depth exploration of the findings.

Aside from visual intuitiveness the 3D representation also provided analytical advantages for quantitative characterization of behavioral dynamics. Specifically, by applying Graph-Theory algorithms to these 3D-ML networks, we characterized shortest paths connecting EMA variables across consecutive temporal layers. The 3D trajectory of such shortest-paths could be decomposed in 4 sets of XYZ coordinates of its TL1-starting-node, TL1-exit-node, TL2-entry-node and TL2-ending-node. We then applied k-means clustering to these coordinates in order to dissect “bundles” of behavioral pathways that shared similar 3D trajectories, reflecting similar role in mediating dynamic transitions between behavioral states. This approach was conceptually inspired by the Neuroimaging Tractography analysis dissecting white-matter bundles pathways based on 3D water diffusion direction in axonal tracts[79]. Moreover, Behavioral Diffusion Analysis (BDA) provided a micro-scale characterization of the contribution of individual variables to distinct behavioral bundles, which we estimated quantitatively as the average 3D diffusion direction of paths that traversed each network node. As such, Behavioral-Tractography allows for a flexible navigation of behavioral dynamics from both a large-scale and detailed symptom-level perspective.

### Correlation between SIPS clinical patterns and dynamic interaction between EMA items in daily life

Next, we explored whether Behavioral-Tractography could identify differences in EMA psychological dynamics corresponding to the differential clinical patterns described by the SIPS. We firstly investigated how SIPS-PCA dimensions moderated the strength of associations between individual pairs of EMA variables, which was achieved by adding each subject’s SIPS-PCA score to the MMLR and focusing on the interaction term between this subject-specific SIPS-PCA score and the strength of association between EMA variables. The model was formalized as follows: *Y _ij_*=*β*_0_ + *β*_1_ *X _ij_* + *β*_2_ *S _j_* + *β*_3_ ( *X _ij_*∗*S _j_* )+*u_j_* +*ε_ij_* where *Y _ij_* is the outcome variable for subject *j* at observation *i, X_ij_* is the predictor variable for subject *j* at observation *i, S _j_* is subject-specific score (SIPS-PCA dimension score), same for all observations of subject j, *X _ij_*∗*S _j_* is interaction term between the EMA predictor and the subject-specific score, *β*_0_ is the fixed intercept, *β*_1_ is the fixed slope, *β*_2_ fixed effect for subject-specific score, *β*_3_ effect of the interaction between the EMA predictor and the subject-specific score, *u _j_* is the random intercept per subject, *ε_ij_* is the residual error term. We repeated this analysis for both SIPS-PCA dimensions and employed the 3D-MLN view to graphically represent both contemporaneous and time-lagged connections that were significantly moderated by SIPS-PCA scores after Benjamini Hochberg multiple comparisons correction [78]. See Figure 3.

We then employed Behavioral-Tractography to provide a meta-scale representation of how SIPS-PCA dimensions influenced dynamic behavioral interactions between EMA variables. First, subjects were grouped into clusters in SIPS-PCA space with k-means clustering as described previously. Separate networks were constructed in each sub-population for which we computed population-specific shortest-paths connecting past to future behavioral variables across temporal layers. We then applied a modified k-means clustering procedure to pathway coordinates measured in both sub-populations (Start-TL1, Exit-TL1-Population-A, Exit-TL1-Population-B, Entry-TL2-Population-A, Entry-TL2-Population-B, End-TL2). The resulting bundles connected a shared set of TL1 to TL2 variables across populations, while still allowing for potential population-specific differences in behavioral trajectories. Such differences in 3D trajectories would suggest qualitative differences in dynamic behavioral interactions between subsets of EMA variables. To assess significance, we then compared these differences in BT-bundle trajectories against a null distribution obtained by permuting subjects randomly across populations.

## Data Availability

All data produced are available online at https://github.com/andreaimparato/Behavioral-Tractography-Toolbox/

## References

1. Kahn, R.S., et al., Schizophrenia. Nat Rev Dis Primers, 2015. 1: p. 15067.

2. Tandon, R., et al., Definition and description of schizophrenia in the DSM-5. Schizophr Res, 2013. 150(1): p. 3–10.

3. Howes, O.D., B.R. Bukala, and K. Beck, Schizophrenia: from neurochemistry to circuits, symptoms and treatments. Nat Rev Neurol, 2024. 20(1): p. 22–35.

4. Galderisi, S., A. Farden, and S. Kaiser, Dissecting negative symptoms of schizophrenia: History, assessment, pathophysiological mechanisms and treatment. Schizophr Res, 2017. 186: p. 1–2.

5. Millan, M.J., et al., Negative symptoms of schizophrenia: clinical characteristics, pathophysiological substrates, experimental models and prospects for improved treatment. Eur Neuropsychopharmacol, 2014. 24(5): p. 645–92.

6. Stiekema, A.P., et al., Confirmatory Factor Analysis and Differential Relationships of the Two Subdomains of Negative Symptoms in Chronically Ill Psychotic Patients. PLoS One, 2016. 11(2): p. e0149785.

7. Galderisi, S., E. Merlotti, and A. Mucci, Neurobiological background of negative symptoms. Eur Arch Psychiatry Clin Neurosci, 2015. 265(7): p. 543–58.

8. Carpenter, W.T., Jr., D.W. Heinrichs, and L.D. Alphs, Treatment of negative symptoms. Schizophr Bull, 1985. 11(3): p. 440–52.

9. Kirschner, M., A. Aleman, and S. Kaiser, Secondary negative symptoms - A review of mechanisms, assessment and treatment. Schizophr Res, 2017. 186: p. 29–38.

10. McCutcheon, R.A., A. Abi-Dargham, and O.D. Howes, *Schizophrenia, Dopa*mine *and the Striatum: From Biology to Symptoms*. Trends Neurosci, 2019. 42(3): p. 205–220.

11. Takeuchi, H., et al., Effects of risperidone and olanzapine dose reduction on cognitive function in stable patients with schizophrenia: an open-label, randomized, controlled, pilot study. Schizophr Bull, 2013. 39(5): p. 993–8.

12. Marder, S.R. and D. Umbricht, Negative symptoms in schizophrenia: Newly emerging measurements, pathways, and treatments. Schizophr Res, 2023. 258: p. 71–77.

13. Kirkpatrick, B., L. Luther, and G.P. Strauss, Negative symptoms in the clinic: we treat what we can describe. Br J Psychiatry, 2023. 223(1): p. 271–272.

14. Daniel, D.G., et al., Remote Assessment of Negative Symptoms of Schizophrenia. Schizophr Bull Open, 2023. 4(1): p. sgad001.

15. Myin-Germeys, I., et al., Experience sampling research in psychopathology: opening the black box of daily life. Psychol Med, 2009. 39(9): p. 1533–47.

16. van Os, J., et al., Beyond DSM and ICD: introducing "precision diagnosis" for psychiatry using momentary assessment technology. World Psychiatry, 2013. 12(2): p. 113–7.

17. Myin-Germeys, I., et al., Emotional reactivity to daily life stress in psychosis. Arch Gen Psychiatry, 2001. 58(12): p. 1137–44.

18. Oorschot, M., et al., Mobile assessment in schizophrenia: a data-driven momentary approach. Schizophr Bull, 2012. 38(3): p. 405–13.

19. Bell, V., et al., Do loneliness and social exclusion breed paranoia? An experience sampling investigation across the psychosis continuum. Schizophr Res Cogn, 2023. 33: p. 100282.

20. Luther, L., et al., Negative symptoms in schizophrenia differ across environmental contexts in daily life. J Psychiatr Res, 2023. 161: p. 10–18.

21. Strauss, G.P., et al., Network analysis of discrete emotional states measured via ecological momentary assessment in schizophrenia. Eur Arch Psychiatry Clin Neurosci, 2023. 273(8): p. 1863–1871.

22. Luther, L., et al., Environmental context predicts state fluctuations in negative symptoms in youth at clinical high risk for psychosis. Psychol Med, 2023. 53(16): p. 7609–7618.

23. Paetzold, I., et al., Momentary Manifestations of Negative Symptoms as Predictors of Clinical Outcomes in People at High Risk for Psychosis: Experience Sampling Study. JMIR Ment Health, 2021. 8(11): p. e30309.

24. Hermans, K., et al., Elucidating negative symptoms in the daily life of individuals in the early stages of psychosis. Psychol Med, 2021. 51(15): p. 2599–2609.

25. Myin-Germeys, I., et al., Experience sampling methodology in mental health research: new insights and technical developments. World Psychiatry, 2018. 17(2): p. 123–132.

26. Fava, G.A., C. Ruini, and C. Rafanelli, Psychometric theory is an obstacle to the progress of clinical research. Psychother Psychosom, 2004. 73(3): p. 145–8.

27. Beltz, A.M., et al., Bridging the Nomothetic and Idiographic Approaches to the Analysis of Clinical Data. Assessment, 2016. 23(4): p. 447–458.

28. Molenaar, P.C.M. and C.G. Campbell, The New Person-Specific Paradigm in Psychology. Current Directions in Psychological Science, 2009. 18(2): p. 112–117.

29. Alanis-Lobato, G., P. Mier, and M.A. Andrade-Navarro, Efficient embedding of complex networks to hyperbolic space via their Laplacian. Sci Rep, 2016. 6: p. 30108.

30. McDonald-McGinn, D.M., et al., 22q11.2 deletion syndrome. Nat Rev Dis Primers, 2015. 1: p. 15071.

31. Schneider, M., et al., Psychiatric disorders from childhood to adulthood in 22q11.2 deletion syndrome: results from the International Consortium on Brain and Behavior in 22q11.2 Deletion Syndrome. Am J Psychiatry, 2014. 171(6): p. 627–39.

32. Sandini, C., et al., More than the sum of their parts: A Behavioral Tractography analysis of distress wellbeing and context reveals dynamic signatures of psychosis and autism. medRxiv, 2025: p. 2025.07.09.25330999.

33. Thornton, M.A. and D.I. Tamir, People represent mental states in terms of rationality, social impact, and valence: Validating the 3d Mind Model. Cortex, 2020. 125: p. 44–59.

34. Thornton, M.A., M.E. Weaverdyck, and D.I. Tamir, The Social Brain Automatically Predicts Others’ Future Mental States. J Neurosci, 2019. 39(1): p. 140–148.

35. Thornton, M.A., M.E. Weaverdyck, and D.I. Tamir, The brain represents people as the mental states they habitually experience. Nat Commun, 2019. 10(1): p. 2291.

36. Thornton, M.A. and D.I. Tamir, Mental models accurately predict emotion transitions. Proc Natl Acad Sci U S A, 2017. 114(23): p. 5982–5987.

37. Tamir, D.I. and M.A. Thornton, Modeling the Predictive Social Mind. Trends Cogn Sci, 2018. 22(3): p. 201–212.

38. Pisani, S., et al., The relationship between alexithymia and theory of mind: A systematic review. Neurosci Biobehav Rev, 2021. 131: p. 497–524.

39. Hitchcock, P.F., E.I. Fried, and M.J. Frank, Computational Psychiatry Needs Time and Context. Annu Rev Psychol, 2022. 73: p. 243–270.

40. Epskamp, S., et al., Personalized Network Modeling in Psychopathology: The Importance of Contemporaneous and Temporal Connections. Clin Psychol Sci, 2018. 6(3): p. 416–427.

41. Mosolov, S.N. and P.A. Yaltonskaya, Primary and Secondary Negative Symptoms in Schizophrenia. Front Psychiatry, 2021. 12: p. 766692.

42. Schneider, M., et al., Preliminary structure and predictive value of attenuated negative symptoms in 22q11.2 deletion syndrome. Psychiatry Res, 2012. 196(2-3): p. 277–84.

43. Schneider, M., et al., Predominant negative symptoms in 22q11.2 deletion syndrome and their associations with cognitive functioning and functional outcome. J Psychiatr Res, 2014. 48(1): p. 86–93.

44. !!! INVALID CITATION !!! [14].

45. Rauthmann, J.F., K.T. Horstmann, and R.A. Sherman, Do Self-Reported Traits and Aggregated States Capture the Same Thing? A Nomological Perspective on Trait-State Homomorphy. Social Psychological and Personality Science, 2018. 10(5): p. 596–611.

46. van Os, J. and U. Reininghaus, Psychosis as a transdiagnostic and extended phenotype in the general population. World Psychiatry, 2016. 15(2): p. 118–24.

47. Sass, L. and G. Byrom, Phenomenological and neurocognitive perspectives on delusions: A critical overview. World Psychiatry, 2015. 14(2): p. 164–73.

48. Kapur, S., Psychosis as a state of aberrant salience: a framework linking biology, phenomenology, and pharmacology in schizophrenia. Am J Psychiatry, 2003. 160(1): p. 13–23.

49. Fletcher, P.C. and C.D. Frith, Perceiving is believing: a Bayesian approach to explaining the positive symptoms of schizophrenia. Nat Rev Neurosci, 2009. 10(1): p. 48–58.

50. Angeletos Chrysaitis, N. and P. Series, 10 years of Bayesian theories of autism: A comprehensive review. Neurosci Biobehav Rev, 2023. 145: p. 105022.

51. Noel, J.P., et al., Aberrant causal inference and presence of a compensatory mechanism in autism spectrum disorder. Elife, 2022. 11.

52. Palmer, C.J., R.P. Lawson, and J. Hohwy, Bayesian approaches to autism: Towards volatility, action, and behavior. Psychol Bull, 2017. 143(5): p. 521–542.

53. Friston, K., The Bayesian Savant. Biol Psychiatry, 2016. 80(2): p. 87–89.

54. Mucci, A., et al., Primary and persistent negative symptoms: Concepts, assessments and neurobiological bases. Schizophr Res, 2017. 186: p. 19–28.

55. Boonstra, N., et al., Duration of untreated psychosis and negative symptoms--a systematic review and meta-analysis of individual patient data. Schizophr Res, 2012. 142(1-3): p. 12–9.

56. Mucci, A., et al., Assessment of Negative Symptoms in Schizophrenia: From the Consensus Conference-Derived Scales to Remote Digital Phenotyping. Brain Sci, 2025. 15(1).

57. Misiak, B., et al., Does social isolation predict the emergence of psychotic-like experiences? Results from the experience sampling method study. Compr Psychiatry, 2024. 135: p. 152521.

58. Strauss, G.P., et al., Markov chain analysis indicates that positive and negative emotions have abnormal temporal interactions during daily life in schizophrenia. J Psychiatr Res, 2023. 164: p. 344–349.

59. Gerritsen, C., et al., Stress precedes negative symptom exacerbations in clinical high risk and early psychosis: A time-lagged experience sampling study. Schizophr Res, 2019. 210: p. 52–58.

60. Formisano, R., et al., The dopamine membrane transporter plays an active modulatory role in synaptic dopamine homeostasis. J Neurosci Res, 2022. 100(8): p. 1551–1559.

61. Fischer, K.D., L.A. Knackstedt, and P.A. Rosenberg, Glutamate homeostasis and dopamine signaling: Implications for psychostimulant addiction behavior. Neurochem Int, 2021. 144: p. 104896.

62. Best, J.A., H.F. Nijhout, and M.C. Reed, Homeostatic mechanisms in dopamine synthesis and release: a mathematical model. Theor Biol Med Model, 2009. 6: p. 21.

63. Koob, G.F. and M. Le Moal, Drug abuse: hedonic homeostatic dysregulation. Science, 1997. 278(5335): p. 52–8.

64. Tiego, J., et al., Precision behavioral phenotyping as a strategy for uncovering the biological correlates of psychopathology. Nature Mental Health, 2023. 1(5): p. 304–315.

65. Wright, A.G.C. and J. Zimmermann, Applied ambulatory assessment: Integrating idiographic and nomothetic principles of measurement. Psychol Assess, 2019. 31(12): p. 1467–1480.

66. Maj, M., The need for a conceptual framework in psychiatry acknowledging complexity while avoiding defeatism. World Psychiatry, 2016. 15(1): p. 1–2.

67. Tyrer, P., Dimensions fit the data, but can clinicians fit the dimensions? World Psychiatry, 2018. 17(3): p. 295–296.

68. Fava, G.A., C. Rafanelli, and E. Tomba, The Clinical Process in Psychiatry. The Journal of Clinical Psychiatry, 2012. 73(02): p. 177–184.

69. Reichert, M., et al., Ambulatory assessment for precision psychiatry: Foundations, current developments and future avenues. Exp Neurol, 2021. 345: p. 113807.

70. Sandini, C., et al., Characterization and prediction of clinical pathways of vulnerability to psychosis through graph signal processing. Elife, 2021. 10.

71. Mancini, V., et al., Positive psychotic symptoms are associated with divergent developmental trajectories of hippocampal volume during late adolescence in patients with 22q11DS. Mol Psychiatry, 2019.

72. Reich, W., Diagnostic Interview for Children and Adolescents (DICA). Journal of the American Academy of Child & Adolescent Psychiatry, 2000. 39(1): p. 59–66.

73. Kaufman, J., et al., Schedule for Affective Disorders and Schizophrenia for School-Age Children-Present and Lifetime Version (K-SADS-PL): Initial Reliability and Validity Data. Journal of the American Academy of Child & Adolescent Psychiatry, 1997. 36(7): p. 980–988.

74. M. First, R.S., J. Williams, *Structured Clinical Interview for the DSM-IV Axis I Disorders (SCID I).* Biometrics Research, New York State Psychiatric Institute, New York, 1996.

75. M. B. First, et al., Structured clinical interview for DSM-5 disorders: SCID-5-CV clinician version, A.P.A. Publishing, Editor. 2016.

76. Miller, T.J., et al., Prodromal assessment with the structured interview for prodromal syndromes and the scale of prodromal symptoms: predictive validity, interrater reliability, and training to reliability. Schizophr Bull, 2003. 29(4): p. 703–15.

77. Feller, C., et al., Psychotic experiences in daily-life in adolescents and young adults with 22q11.2 deletion syndrome: An Ecological Momentary Assessment study. Schizophr Res, 2021. 238: p. 54–61.

78. Genovese, C.R., N.A. Lazar, and T. Nichols, Thresholding of statistical maps in functional neuroimaging using the false discovery rate. Neuroimage, 2002. 15(4): p. 870–8.

79. Wandell, B.A., Clarifying Human White Matter. Annu Rev Neurosci, 2016. 39: p. 103–28.

