## Supplementary_Material for "Shedding light on the dynamic interplay of positive and negative symptoms of psychosis with Behavioral Tractography"

### SUPPLEMENTARY MATERIAL INDEX


- **Supplementary Table 1. Behavioral Diffusion Analysis (BDA)**
- **Supplementary Analysis 1: Replication of Behavioral Tractography and Diffusion Analysis in 22q11DS and Typically Developing Sample**
- **EMA Protocol**
- **References**

**Supplementary Table 1**. Behavioral Diffusion Analysis (BDA). Detailed quantitative comparison of BDA-derived trajectories across samples, highlighting how specific symptoms contribute to behavioral dynamics. The columns (CL1–CL9) represent distinct bundles derived from the Behavioral-Tractography analysis. The table is organized into the two SIPS-PCA derived dimensions: High-/Low-Symptom-Intensity Dimension and Predominant Positive/Negative Symptom Dimension.

The first row (BT-P-values) of the table presents the global p-values for the differences in BT-bundle trajectories between samples, tested against a null distribution obtained by randomly permuting subjects across populations. The subsequent rows display the BDA analysis p-values, which are organized by Temporal Layers 1 and 2. These reflect the statistical significance of specific symptom contributions to behavioral dynamics within each bundle and temporal layer.

| | High-/ Low-Symptom-Intensity Dimension | | | | | | | | | | Predominant Positive- negative Symptom Dimension | | | | | | | | | |
|---|---|---|---|---|---|---|---|---|---|---|---|---|---|---|---|---|---|---|---|---|
| | BT-P-values | CL1 0.86 | CL2 0.07 | CL3 0.99 | CL4 0.29 | CL5 0.76 | CL6 0.28 | CL7 0.03 | CL8 0.08 | CL9 0.00 | BT-P-values | **CL1 0.00** | **CL2 0.24** | CL3 0.36 | CL4 0.21 | CL5 0.05 | CL6 0.26 | CL7 0.00 | **CL8 0.00** | **CL9 0.00** |
| | EMA Symptoms | CL1 | CL2 | CL3 | CL4 | CL5 | CL6 | CL7 | CL8 | CL9 | EMA Symptoms | CL1 | CL2 | CL3 | CL4 | CL5 | CL6 | CL7 | CL8 | CL9 |
| TEMPORAL LAYER 1 | Lack Relaxation | 0.84 | 1.00 | 0.81 | 0.99 | 0.75 | 0.42 | 0.63 | 0.56 | 0.00 | Lack Relaxation | 0.82 | 1.00 | 0.55 | 1.00 | 0.05 | 0.91 | 1.00 | 0.7 | 0.45 |
| | Loneliness | 1.00 | 0.70 | 0.73 | 0.15 | 1.00 | 0.88 | 1.00 | 0.39 | 0.10 | Loneliness | 1.00 | 0.21 | 1.00 | 1.00 | 0.58 | 1.00 | 0.22 | 0.3 | 0.08 |
| | Anxiety | 0.90 | 0.45 | 0.89 | 0.65 | 0.81 | 1.00 | 1.00 | 0.99 | 0.03 | Anxiety | 0.65 | 1.00 | 0.91 | 1.00 | 0.39 | 1.00 | 1.00 | 0.9 | 0.26 |
| | Lack Happiness | 0.47 | 0.01 | 0.82 | 0.25 | 0.24 | 0.04 | 0.31 | 1.00 | 0.33 | Lack Happiness | 0.02 | 0.20 | 0.13 | 0.88 | 0.27 | 0.33 | 0.76 | 1.0 | 0.16 |
| | Irritation | 0.30 | 1.00 | 0.12 | 0.13 | 0.30 | 1.00 | 0.28 | 0.76 | 0.25 | Irritation | 0.76 | 1.00 | 0.20 | 1.00 | 0.48 | 1.00 | 1.00 | 0.2 | 0.30 |
| | Sensory Issue | 0.97 | 0.23 | 0.96 | 0.32 | 1.00 | 1.00 | 1.00 | 0.97 | 0.21 | Sensory Issue | 0.77 | 0.90 | 1.00 | 1.00 | 0.05 | 0.22 | 0.59 | 0.0 | 0.37 |
| | Lack Excitement | 0.18 | 1.00 | 0.07 | 1.00 | 0.62 | 0.59 | 0.45 | 1.00 | 0.06 | Lack Excitement | 1.00 | 1.00 | 0.77 | 0.84 | 0.00 | 0.02 | 0.00 | 1.0 | 0.09 |
| | Sadness | 0.88 | 0.00 | 0.88 | 0.32 | 0.45 | 0.15 | 1.00 | 0.98 | 0.07 | Sadness | 0.07 | 1.00 | 0.64 | 1.00 | 0.23 | 1.00 | 1.00 | 0.0 | 0.03 |
| | Lack Confidence | 0.97 | 1.00 | 0.08 | 0.12 | 0.93 | 1.00 | 1.00 | 1.00 | 1.00 | Lack Confidence | 0.30 | 1.00 | 0.64 | 1.00 | 0.35 | 1.00 | 1.00 | 1.0 | 0.56 |
| | Feeling Rejected | 0.31 | 0.90 | 0.75 | 1.00 | 0.93 | 1.00 | 1.00 | 0.07 | 0.28 | Feeling Rejected | 0.62 | 0.51 | 1.00 | 1.00 | 0.96 | 0.33 | 1.00 | 0.8 | 0.28 |
| | Feeling Unsafe | 1.00 | 0.58 | 0.52 | 1.00 | 0.88 | 1.00 | 1.00 | 0.22 | 0.46 | Feeling Unsafe | 0.11 | 0.07 | 1.00 | 1.00 | 0.28 | 1.00 | 0.36 | 0.8 | 0.39 |
| | Confusion | 1.00 | 0.18 | 0.95 | 0.71 | 1.00 | 1.00 | 1.00 | 0.06 | 0.95 | Confusion | 0.06 | 0.11 | 1.00 | 1.00 | 0.35 | 1.00 | 0.75 | 0.3 | 0.91 |
| | Hallucinations | 1.00 | 0.00 | 0.72 | 0.84 | 0.65 | 1.00 | 1.00 | 0.33 | 0.22 | Hallucinations | 0.64 | 0.98 | 1.00 | 1.00 | 0.38 | 1.00 | 0.96 | 0.5 | 0.36 |
| | Feeling Tired | 1.00 | 0.85 | 0.50 | 0.15 | 1.00 | 0.33 | 0.05 | 0.44 | 0.40 | Feeling Tired | 0.08 | 0.00 | 1.00 | 0.61 | 0.05 | 0.91 | 0.74 | 0.3 | 0.14 |
| | Lack Motivation | 1.00 | 0.09 | 0.65 | 0.14 | 1.00 | 0.41 | 0.65 | 1.00 | 1.00 | Lack Motivation | 1.00 | 1.00 | 1.00 | 0.12 | 1.00 | 0.08 | 0.53 | 1.0 | 1.00 |
| | Lack Physical Activity | 1.00 | 0.68 | 1.00 | 0.03 | 1.00 | 0.03 | 0.33 | 1.00 | 1.00 | Lack Physical Activity | 1.00 | 1.00 | 1.00 | 0.38 | 1.00 | 0.53 | 0.46 | 1.0 | 0.06 |
| | Finding Activity Difficult | 1.00 | 0.94 | 1.00 | 0.75 | 1.00 | 0.91 | 0.49 | 0.38 | 1.00 | Finding Activity Difficult | 0.75 | 1.00 | 1.00 | 0.89 | 1.00 | 0.95 | 0.38 | 0.6 | 1.00 |
| | Lack Enjoying Activity | 1.00 | 1.00 | 1.00 | 0.77 | 0.17 | 0.84 | 0.58 | 1.00 | 1.00 | Lack Enjoying Activity | 0.37 | 1.00 | 1.00 | 0.17 | 1.00 | 0.47 | 0.05 | 0.41 | 0.05 |
| | Lack Concentration | 1.00 | 0.77 | 1.00 | 0.52 | 1.00 | 0.06 | 0.12 | 1.00 | 1.00 | Lack Concentration | 1.00 | 1.00 | 1.00 | 0.01 | 0.64 | 0.12 | 1.00 | 1.00 | 1.00 |
| | Being Alone | 1.00 | 0.07 | 1.00 | 0.01 | 1.00 | 0.24 | 1.00 | 0.10 | 1.00 | Being Alone | 1.00 | 0.40 | 1.00 | 1.00 | 0.37 | 1.00 | 0.49 | 0.20 | 0.42 |
| | Symptoms | CL1 | CL2 | CL3 | CL4 | CL5 | CL6 | CL7 | CL8 | CL9 | Symptoms | CL1 | CL2 | CL3 | CL4 | CL5 | CL6 | CL7 | CL8 | CL9 |
| TEMPORAL LAYER 2 | Lack Relaxation | 0.54 | 1.00 | 0.76 | 0.21 | 1.00 | 0.50 | 0.60 | 0.44 | 0.06 | Lack Relaxation | 0.90 | 1.00 | 0.10 | 0.31 | 1.00 | 0.00 | 0.00 | 0.00 | 0.99 |
| | Loneliness | 1.00 | 0.56 | 1.00 | 0.90 | 0.00 | 0.36 | 0.00 | 0.23 | 1.00 | Loneliness | 0.62 | 0.63 | 0.03 | 0.00 | 0.10 | 0.15 | 0.63 | 1.00 | 1.00 |
| | Anxiety | 0.56 | 0.64 | 0.95 | 0.47 | 0.52 | 0.04 | 0.18 | 0.33 | 0.40 | Anxiety | 0.44 | 1.00 | 0.48 | 1.00 | 0.96 | 0.06 | 0.06 | 0.13 | 1.00 |
| | Lack Happiness | 0.89 | 1.00 | 0.97 | 0.34 | 0.99 | 0.81 | 0.98 | 0.40 | 0.91 | Lack Happiness | 0.35 | 1.00 | 0.72 | 0.03 | 1.00 | 0.19 | 0.36 | 0.07 | 1.00 |
| | Irritation | 0.13 | 0.15 | 1.00 | 0.09 | 1.00 | 0.11 | 0.09 | 0.14 | 0.13 | Irritation | 0.78 | 1.00 | 1.00 | 0.08 | 0.12 | 0.03 | 0.00 | 0.25 | 1.00 |
| | Sensory Issue | 1.00 | 0.14 | 1.00 | 0.63 | 0.00 | 1.00 | 0.00 | 0.98 | 0.98 | Sensory Issue | 1.00 | 0.14 | 0.00 | 1.00 | 0.11 | 0.04 | 0.00 | 0.28 | 1.00 |
| | Lack Excitement | 1.00 | 1.00 | 0.11 | 0.21 | 1.00 | 0.05 | 1.00 | 0.00 | 0.04 | Lack Excitement | 1.00 | 1.00 | 0.77 | 0.51 | 1.00 | 0.66 | 1.00 | 0.00 | 0.00 |
| | Sadness | 0.92 | 0.16 | 1.00 | 1.00 | 0.28 | 0.16 | 0.07 | 0.00 | 0.93 | Sadness | 0.03 | 1.00 | 0.02 | 0.01 | 0.09 | 0.00 | 1.00 | 0.85 | 1.00 |
| | Lack Confidence | 0.88 | 1.00 | 1.00 | 0.99 | 1.00 | 0.00 | 0.00 | 0.09 | 0.10 | Lack Confidence | 0.49 | 1.00 | 0.27 | 0.06 | 1.00 | 0.00 | 1.00 | 0.02 | 1.00 |
| | Feeling Rejected | 0.96 | 0.94 | 1.00 | 1.00 | 0.96 | 0.09 | 0.07 | 0.92 | 1.00 | Feeling Rejected | 0.20 | 0.41 | 0.07 | 1.00 | 1.00 | 0.02 | 0.35 | 0.15 | 1.00 |
| | Feeling Unsafe | 1.00 | 0.00 | 1.00 | 1.00 | 0.02 | 1.00 | 0.15 | 1.00 | 1.00 | Feeling Unsafe | 0.26 | 0.81 | 1.00 | 1.00 | 0.30 | 0.35 | 0.00 | 0.57 | 1.00 |
| | Confusion | 1.00 | 0.89 | 1.00 | 0.71 | 0.10 | 1.00 | 0.10 | 0.99 | 1.00 | Confusion | 0.12 | 0.76 | 1.00 | 1.00 | 0.01 | 0.00 | 0.00 | 1.00 | 1.00 |
| | Hallucinations | 1.00 | 0.27 | 1.00 | 0.84 | 0.00 | 1.00 | 0.09 | 0.16 | 1.00 | Hallucinations | 1.00 | 0.99 | 1.00 | 1.00 | 0.00 | 0.00 | 0.00 | 0.89 | 1.00 |
| | Feeling Tired | 1.00 | 0.75 | 0.51 | 0.91 | 1.00 | 1.00 | 0.55 | 0.93 | 0.11 | Feeling Tired | 1.00 | 0.86 | 1.00 | 0.55 | 0.06 | 0.28 | 0.33 | 0.62 | 0.21 |
| | Lack Motivation | 1.00 | 1.00 | 0.22 | 0.27 | 1.00 | 1.00 | 0.99 | 0.12 | 0.04 | Lack Motivation | 1.00 | 1.00 | 0.00 | 0.60 | 1.00 | 1.00 | 1.00 | 0.21 | 0.05 |
| | Lack Physical Activity | 1.00 | 1.00 | 0.13 | 0.20 | 1.00 | 1.00 | 1.00 | 1.00 | 1.00 | Lack Physical Activity | 1.00 | 1.00 | 0.00 | 0.13 | 1.00 | 1.00 | 0.95 | 1.00 | 0.00 |
| | Finding Activity Difficult | 1.00 | 1.00 | 0.01 | 0.99 | 1.00 | 1.00 | 0.87 | 0.52 | 0.00 | Finding Activity Difficult | 0.01 | 1.00 | 0.00 | 0.32 | 1.00 | 1.00 | 0.99 | 0.00 | 0.00 |
| | Lack Enjoying Activity | 1.00 | 1.00 | 0.03 | 0.26 | 1.00 | 0.99 | 0.96 | 0.05 | 0.00 | Lack Enjoying Activity | 0.01 | 1.00 | 0.03 | 0.09 | 1.00 | 1.00 | 0.67 | 0.00 | 0.11 |
| | Lack Concentration | 1.00 | 1.00 | 0.11 | 0.06 | 1.00 | 1.00 | 0.99 | 0.03 | 0.03 | Lack Concentration | 1.00 | 1.00 | 1.00 | 0.94 | 1.00 | 1.00 | 1.00 | 0.00 | 0.00 |
| | Being Alone | 1.00 | 0.69 | 1.00 | 0.54 | 1.00 | 0.66 | 0.03 | 1.00 | 1.00 | Being Alone | 1.00 | 0.75 | 0.00 | 0.00 | 0.49 | 1.00 | 0.82 | 1.00 | 1.00 |

### Supplementary Analysis 1: Replication of Behavioral Tractography and Diffusion Analysis in 22q11DS and Typically Developing Sample

### Methods

To assess the reproducibility of Behavioral-Tractography and Behavioral Diffusion Analysis (BDA) results, we performed a replication analysis comparing the 22q11DS sample with findings from our previous study on a typically developing population (TD-D) [1]. The goal was to determine whether the composition and average trajectories of Behavioral-Tractography-Bundles observed in the 22q11DS cohort replicate the psychological-contextual interaction patterns previously identified in TD samples. The TD-D sample included 53 typically developing individuals (26 females, mean age = 18.5. SD = 3.7) recruited from the Geneva community and siblings of 22q11DS carriers. All participants had a sufficient command of French, provided written consent and met strict inclusion criteria (excluded individuals with premature birth, a first-degree relative with a developmental disorder (except for a de novo 22q11.2 deletion), or a history of psychiatric, neurological, or learning disorders).
First, we assessed consistency by comparing k-means clustering results of 3D psychological-contextual trajectories in both samples and by correlating the mean trajectory of SP pathways within each Behavioral-Tractography bundle across samples. Finally, we validated Dynamic Behavioral Centrality (DBC) and BDA by correlating their values at individual network nodes between the TD-D and 22q samples.

**Results**

Replication analyses showed consistency between the 22q11DS and typically developing (TD-D) samples in both Behavioral-Tractography and Behavioral Diffusion Analysis (BDA). As shown in Panel 13, the average 3D coordinates of the nine Behavioral-Tractography-Bundles were correlated across independent samples, with a correlation coefficient of $r=0.87$ ($p<0.001$), indicating reproducibility of bundle composition and trajectories. In Panel 14, BDA trajectories quantified by 8 XY coordinates for each bundle and variable across all 40 network nodes-also showed significant correlation across samples ($r=0.50$, $p<0.001$), supporting the replicability of psychological-contextual interaction patterns. When the analysis was restricted to variables that mediated at least one path in both samples, the correspondence increased ($r=0.80$, $p<0.001$), as indicated by the red regression line. Together, these results indicate that both the composition of Behavioral-Tractography bundles and the underlying BDA trajectories observed in 22q11DS replicate those previously identified in typically developing individuals.

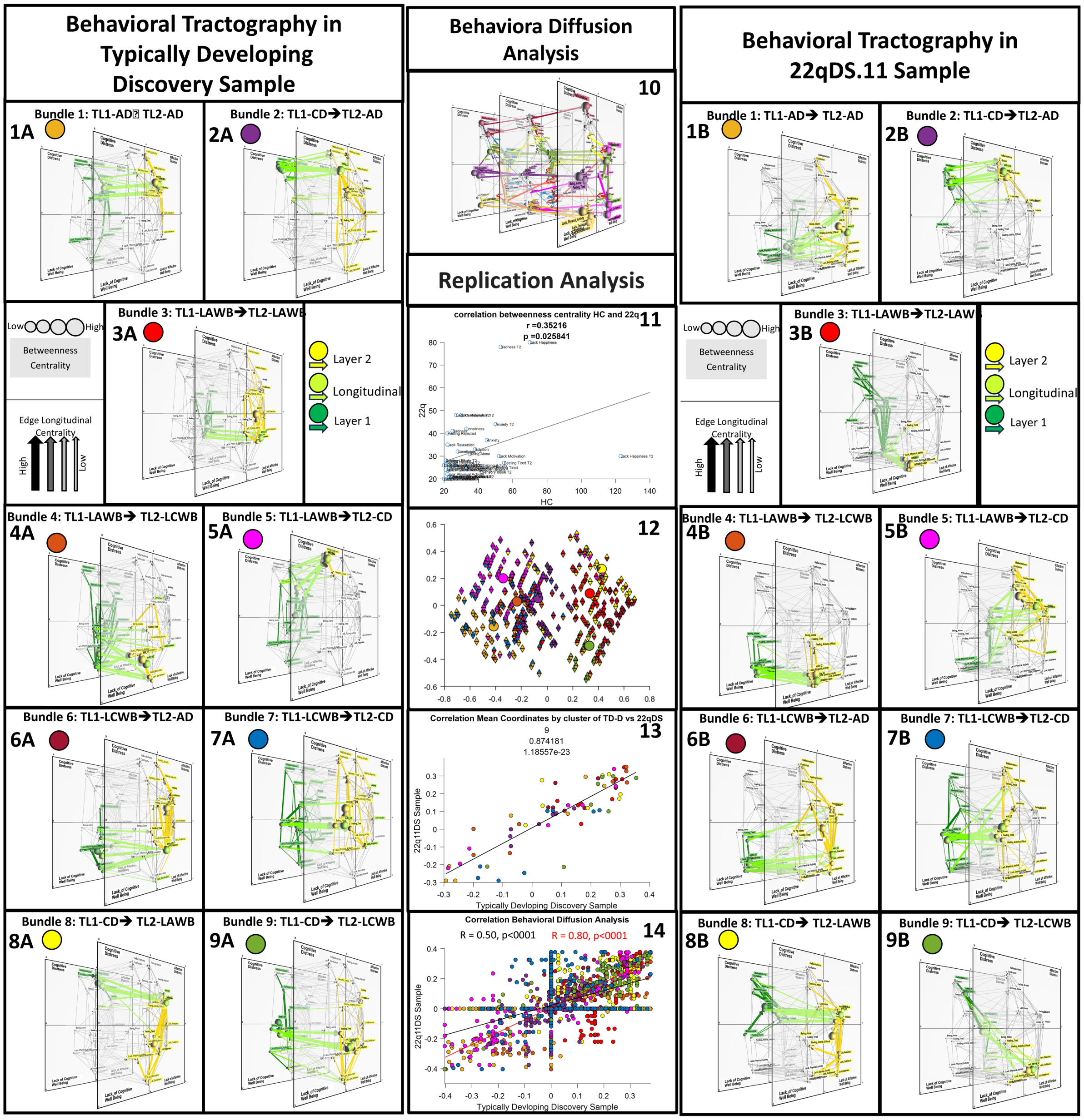


**Supplementary figure 1**: Replication of Behavioral Tractography and Diffusion Analysis in 22q11DS and Typically Developing Sample. **Panels 1A-1B:** Bundle 1: TL1-AD-->TL2-AD. **Panels 2A-2B:** Bundle 2: TL1-CD-->TL2-AD. **Panels 3A-3B:** Bundle 3: TL1-LAWB-->TL2- LAWB. **Panels 4A-4B:** Bundle 4: TL1- LAWB -->TL2-LCWB. **Panels 5A-5B:** Bundle 5: TL1- LAWB -->TL2-CD. **Panels 6A-6B**: Bundle 6: TL1-LCWB-->TL2-AD. **Panels 7A-7B:** Bundle 7: TL1- LCWB -->TL2-CD. **Panels 8A-8B:** Bundle 8: TL1-CD-->TL2-LAWB. **Panels 9A-9B:** Bundle 9: TL1-CD-->TL2-LCWB. Green to Yellow color coding reflects the dynamic progressing of Shortest Paths from TL1 to TL2 (Start-TL1==>Exit-TL1: Dark Green; Exit-TL1==>Entry-TL2: Light Green; Entry-TL2==>End-TL2: Yellow). **Panels 10:** BDA trajectories of variables that disproportionately mediate PC pathways in each Behavioral-Tractography Bundle in TD-D Sample. Color coding of BDA vectors reflects the Behavioral-Tractography bundle that is significantly mediated by the corresponding variables. Variables that act as gateways for subsequent PC states are represented in TL-1 with their respective BDA trajectories connecting TL-1 and TL-2. Variables that act as funnels for previous PC states are represented in TL-3 with their respective BDA trajectories connecting TL-2 and TL-3. **Panel 11:** Replication analysis of Longitudinal Betweenness Centrality (Longi-BC) across TD-D and 22q samples. **Panel 12:** Replication analysis of Behavioral Tractography across TD-D and 22q samples in terms of composition of Behavioral-Tractography Bundles. Each triangle represents a Psychological-Contextual pathway connecting variables across temporal layers. Pathways are plotted in two-dimensional space derived from Principal-Component-Analysis of their 3D trajectory. PCA was performed on the same data-matrix to which K-means was applied to dissect Behavioral-Tractography bundles. which was composed of a line for each pathway each and 8 columns codifying it's 3-D Trajectory (X-Y-Starting-TL1. X-Y-Exit-TL1. X-Y-Entry-TL2. X-Y-End-TL2). Color of triangles represent the attribution of pathways to one of 9 Behavioral-Tractography-Bundles. Upward-pointing triangles represent composing Behavioral-Tractography Bundles in TD-D sample and downward-pointing triangles in 22q sample. **Panel 13:** Replication of average 3D coordinates of the 9 Behavioral-Tractography-Bundles computed in independent TD-D and 22q sample. Each point represents either the x or y value of one of the 4 coordinates for each bundle. **Panel 14:** Replication analysis of BDA trajectories computed for each variable and each Behavioral-Tractography bundle across TD-D and 22q samples. BDA trajectories are codified by 8 XY coordinates and computed across 9 clusters for all 40 network nodes and yielding a total of 2880 points that are correlated across samples as represented by the black regression line. When a given variable did not mediate any PC-path in a given Behavioral-Tractography bundle the corresponding BDA coordinates were set to 0. We also repeated the BDA replication analysis considering only variables that mediated at least one path in the corresponding Behavioral-Tractography-Bundle in both samples, as represented by the red regression line.

**Ema Protocol**

**General Informations**

- 8 notifications per day between 7:30 a.m. and 10:00 p.m. for 6 days
- At least 30 minutes between two notifications
- 15-minute window to respond before the notification disappears
- 3 blocks of questions (mood/symptomatology, context, event)

**EMA Items**

All EMA items were originally written in French and are presented here translated into English. Items shown in bold indicate those selected for analysis in this paper.

- **Mood/Symptoms**
    - **Right now, I feel relaxed (var = Lack Relaxation)**
    - **Right now, I feel lonely (var = Loneliness)**
    - **Right now, I feel anxious (var = Anxiety)**
    - **Right now, I feel happy, joyful (var = Lack Happiness)**
    - **Right now, I feel irritated, angry (var = Irritation)**
    - **Right now, I feel bothered by sensory stimulation (var = Sensory Issue)**
    - **Right now, I feel excited (var = Lack Excitement)**
    - **Right now, I feel sad (var = Sadness)**
    - **Right now, I feel confident (var = Lack Confidence)**
    - **Right now, I feel like others don't like me (var = Feeling Rejected)**
    - **Right now, I feel like I have to stay on guard, that I'm not safe (var = Feeling Unsafe)**
    - **Right now, I feel like my imagination is blending with reality (var = Confusion)**
    - **Right now, I feel like I'm hearing or seeing things that others don't perceive (var = Hallucinations)**
    - **Right now, I feel tired (var = Feeling Tired)**
    - **Right now, I feel like doing many things, I feel motivated (var = Lack Motivation)**
    - **Since the last beep, I have been physically active (var = Lack Physical Activity)**

- **Contexte**
    - What are you doing? (just before the beep) (var = Cur_activ)
    - **This activity is difficult (var = Finding Activity Difficult)**
    - **I am enjoying this activity (var = Lack Enjoying Activity)**
    - **I am focused on this activity (var = Lack Concentration)**
    - Where are you? (juste avant le beep) (var = Location)
    - **Are you alone? (var = Being Alone)**
    - Yes (=1)
        - I would prefer to be with other people (var = Pref_other)
        - I like being alone (var = Likealone)
        - I feel excluded, rejected (var = Feel_exclu)
    - No (=0)
        - Who are you with?
        - I would prefer to be alone (var = Pref_alone)
        - This company is pleasant (var = likecomp)

    - We are doing something together (var = interact)
    - I feel judged by this/these person(s) (var = feel_judged)
    - I am nervous in the presence of this/these person(s) (var = feel_nerv)
- Evénement
    - Now think about the most important event that has happened since the last beep
    - This event was pleasant
    - This event was stressful
    - This event was important
    - Were you alone during this event?
    - Now think about the most important event that will happen in the next hour
    - I am looking forward to this event
    - In which category does this event fall?
    - Will you be alone during this event?

**Response scale:**

1 (not at all), 2 (very slightly), 3 (slightly), 4 (moderately), 5 (strongly), 6 (very strongly), 7 (extremely)

**Context: activity:**
10 (work/school), 20 (household tasks), 30 (eating or drinking), 40 (personal care), 50 (rest/relaxation), 60 (social contact), 65 (online social contact), 70 (sports), 75 (TV, internet), 79 (other leisure), 89 (something else), 0 (nothing)

**Context: location:**
10 (home), 20 (school/work), 30 (at a friend's place), 40 (at a family member's place), 50 (hospital or care facility), 60 (public place), 70 (in transit), 89 (elsewhere)

**Context: with whom:**
10 (person living with me), 20 (person not living with me), 30 (boyfriend/girlfriend, spouse), 40 (friend), 50 (classmate), 60 (healthcare professional), 70 (acquaintance), 80 (pet), 89 (stranger), 99 (no one else)

**Event:**
0 (nothing), 10 (physical activity), 20 (work/school), 30 (leisure), 40 (sleeping), 50 (eating/drinking), 60 (medical activity), 89 (other)


1. Sandini, C., et al., *More than the sum of their parts: A Behavioral Tractography analysis of distress wellbeing and context reveals dynamic signatures of psychosis and autism.* medRxiv, 2025: p. 2025.07.09.25330999.
